# Estimation of Direct and Indirect Polygenic Effects and Gene-Environment Interactions using Polygenic Scores in Case-Parent Trio Studies

**DOI:** 10.1101/2024.10.08.24315066

**Authors:** Ziqiao Wang, Luke Grosvenor, Debashree Ray, Tianyuan Cheng, Ingo Ruczinski, Terri H. Beaty, Heather Volk, Christine Ladd-Acosta, Nilanjan Chatterjee

## Abstract

Family-based studies provide a unique opportunity to characterize genetic risks of diseases in the presence of population structure, assortative mating, and indirect genetic effects. We propose a novel framework, PGS-TRI, for the analysis of polygenic scores (PGS) in case-parent trio studies to estimate the risk of an index condition associated with direct effects of inherited PGS, indirect effects of parental PGS, and gene-environment interactions. Extensive simulation studies demonstrate the robustness of PGS-TRI in the presence of complex population structure and assortative mating. We applied PGS-TRI to multi-ancestry trio studies of autism spectrum disorders (ASD) (N_trio_ = 18,383), deriving transmission-based estimates of risk for the direct effects of established PGS for ASD and other neurocognitive traits PGS across different ancestry groups and along a genetic ancestry continuum. Our analysis also identified significant indirect effects from parental PGS for BMI and several neurocognitive traits on children’s ASD risk. We also employed PGS-TRI in a trio study of European and Asian orofacial clefts (OFCs) (N_trio_ = 1,904), investigating both direct and indirect effect of an established PGS and its interaction with maternal risk factors. Finally, we applied PGS-TRI to investigate the direct and indirect effects of large-scale transcriptome-wide and metabolome-wide traits on ASD and OFCs risks.

## Introduction

Large genome-wide association studies (GWAS) of unrelated individuals have been widely used to derive polygenic scores (PGS) or polygenic risk scores (PRS) for complex traits. While it is standard practice to account for population stratification using genetic principal components, recent studies^1-5^ have demonstrated the potential for overestimating genetic effects due to residual confounding with geographical variations and/or assortative mating in the population. This complicates translational applications and interpretations for PGS across various analyses, including risk predictions and Mendelian randomization analysis. ^6^ Family-based association studies,^7^ which involve estimating genetic effects through within-family comparisons, can protect against such biases when assessing effects of individual genetic variants as well as PGS. Further, family-based studies with parental data can uniquely be used to estimate the indirect effects^8-11^ of parental genetic variation on offspring outcomes, possibly through parental environmental factors. Thus, these PGS could be used to obtain genetic-based evidence of the effect of parental exposures on children’s health outcomes.

To date, there have been limited studies on methodologies estimating PGS effects associated with disease risks through family-based studies. One study^7^ described methods for separating within and between family effects of PGS based on random-effect models to account for family-specific effects. Another study ^11^ pioneered the use of parental genotype data on probands to separate direct and indirect effects of PGS on traits within families. These methods were primarily developed for the analysis of quantitative traits in randomly sampled families and are not suitable for other important study designs including case-parent trios, mother-child dyads, or other family-based study designs where families are ascertained based on affected probands. One recent study^12^ introduced the polygenic transmission disequilibrium test (pTDT) based on case-parent trio designs and detected evidence of polygenic risk for autism spectrum disorders (ASD), irrespective of the presence of high *de novo* variants in probands. The method, however, does not provide estimates of effect-sizes in a suitable risk-scale, a critically important task for many purposes such as for comparisons of risk estimates from population-based studies, estimating causal effects using Mendelian randomization studies, and defining concepts of interactions.

To address these limitations and meet the current needs for the analysis of PGS in family-based studies, we introduce PGS-TRI. This method can be applied to case-parent trio study designs to estimate the risk of a condition in offspring associated with direct (inherited) effects of PGS and its interaction with environmental exposures (PGSxE), and indirect effects of parental PGS.^8, 10^ The method allows disease risk and PGS distribution to vary across families in a flexible manner, making it highly resilient to population stratification and assortative mating. We show that under our modeling framework, the PGS distribution in ascertained families can be derived in a compact form and can be conveniently partitioned into transmission and parental components. Based on this factorization, we present novel methods for estimating the direct effect of PGS and the effect of PGSxE interactions on offspring outcomes using the transmission component, while using a key scale parameter estimate from the parental PGS distribution. Additionally, we show how parental PGS data can be used to derive a simple and highly robust estimator for the difference in indirect effects of maternal and paternal PGS on offspring’s outcomes. We conduct extensive simulation studies to demonstrate the validity and power of PGS-TRI for detecting direct and indirect effects, and PGSxE interactions under the presence of complex population structures and assortative mating.

We applied PGS-TRI to a large multi-ancestry dataset of case-parent autism spectrum disorder (ASD) trios ascertained from the Simons Powering Autism Research for Knowledge (SPARK) consortium.^13^ We derived established PGS for ASD, several neurocognitive traits, and body mass index (BMI), and investigated risk of ASD in the children associated with the direct and indirect effects of these scores. We observed direct effects for these scores of similar patterns and magnitude to those reported in population-based studies. Moreover, we observed significant indirect effects for the scores for BMI and several neurocognitive traits, but not for ASD itself. We further demonstrated that the heterogeneity in ASD PGS effects across ancestry groups can be explained by a continuous attenuation of association strength, reflecting the genetic distance between the target population from the PGS training population. We further applied the proposed method to a trio study of orofacial clefts (OFCs), another developmental disorder known to be highly heritable. The analysis revealed a strong direct effect of an established PGS across European and Asian populations and multiple disease subtypes, but no evidence of any indirect effects. For both ASD and OFCs, we further evaluated PGSxE interactions for several known maternal risk factors. Finally, we explored the effectiveness of PGS-TRI as a discovery tool by analyzing PGS for gene expression and metabolite traits, obtained from the OMICSPRED study,^14^ to investigate their potential direct or indirect effects on the risk of the two conditions.

## Material and Methods

### Modeling Direct Genetic Effects and Gene-Environment Interactions

An overview of the method is presented in Fig.1. We assume PGS for the index condition and other traits of interest can be evaluated for family trio participants, using published meta-data from prior association studies. Our goal is to investigate the association of these PGS with an index condition, such as ASD, using case-parent trios. In a trio, let *PGS*_*C*_, *PGS*_*M*_, and *PGS*_*F*_ denote the PGS values of the child, mother, and father respectively. Additionally, let *D*_*C*_ denote the binary outcome status of the child. We assume *i* = 1, …, *N* families have been sampled following the case-parent trio design. We also assume a set of environmental exposures, denoted by ***E***_***ic***_, has been ascertained for each child across the different families. We assume prospective disease risk of the child in the population follows a log-linear model:

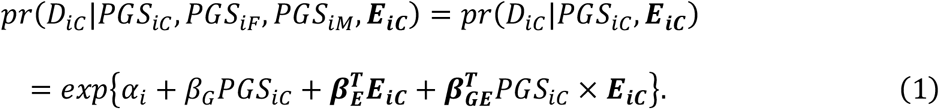

**Figure 1.**
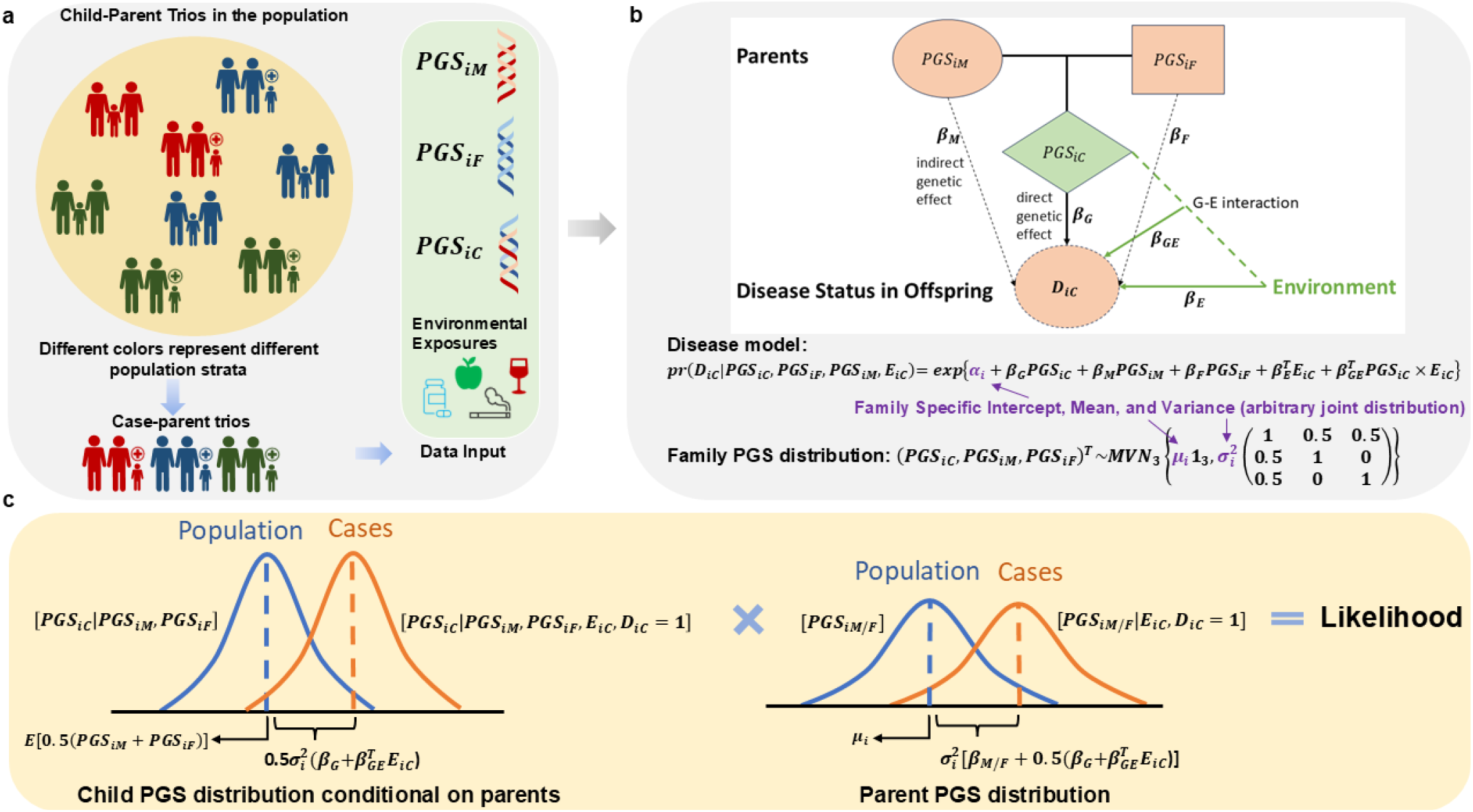
Figure illustrating our PGS-TRI model framework for case-parent trio family study designs. *β*_*G*_, ***β***_***GE***_, ***β***_***E***_: population-level PGS direct genetic effect (*β*_*G*_, direct PGS-E interactions (*β*_*GE*_, and direct environmental effects (*β*_*E*_, associated with offspring’s outcome. *β*_*M*_, *β*_*F*_: mother and father indirect genetic effects associated with the offspring’s outcome risk. *PGS*_*iC*_, *PGS*_*iM*_, *PGS*_*iF*_ PGS values of child, mother, father in famil *i* = 1, …, *N. D*_*iC*_ : outcome status of the offspring in famil *i* = 1, …, *N*.

here, *β*_*G*_ is the direct genetic effect (DE). In (1), we assume no indirect genetic effects, i.e., within each family, the parental PGS values only affect the child’s disease risk mediated through the child’s own PGS value, but we will relax this assumption later (see subsequent sections and Supplemental Notes). Additionally, this model incorporates family-specific intercept terms *α*_*i*_ without any further assumption about their distribution, thereby allowing disease risks to vary arbitrarily across families.

We assume the joint distributions of PGS across families in the underlying population follow trivariate normal distributions of the form:

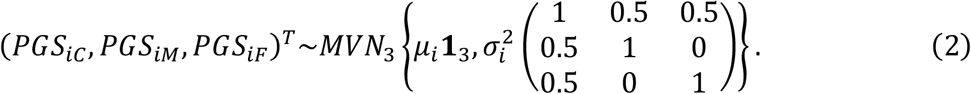

where the correlation of 0.5 between PGS values of individual parents and children follows from Mendel’s law of inheritance. We allow family-specific mean (*μ*_*i*_) and variance (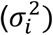) terms without imposing any assumptions regarding their distributions. We assume within a family, PGS for the parents are independently distributed.

We explain here how a model of form (2) can flexibly account for both population structure and assortative mating. We conceptualize a sampling mechanism where for each family sampled, we assume it belongs to a unique subpopulation of “homogeneous” families with distinct genetic ancestry and trait characteristics influencing assortative mating. The number of these subpopulations can be arbitrarily large, making each subpopulation at an extremely fine level. It is assumed random mating occurs within these homogeneous fine-level subpopulations, implying the PGS correlation between partners is 0 within these subpopulations, but not necessarily across them. By allowing arbitrary family-specific parameters for the PGS distribution, the model can accommodate fine-level population structure and the presence of any population-level correlation in PGS values between parents due to assortative mating. Finally, we assume *E*_*iC*_ ⊥ (*PGS*_*iC*_, *PGS*_*iM*_, *PGS*_*iF*_), but allow the distribution ***E***_***iC***_ to remain unspecified. From a population perspective, this assumption can again be viewed as “gene-environment” independence *within* highly homogenous subpopulations, but the model can still accommodate gene-environment correlation at the population-level that may arise due to population substructure and assortative mating.

### Retrospective Likelihood and Parameter Estimation

The retrospective likelihood for the case-parent trio data in each family *i* can be decomposed into an offspring’s (*L*_*iC*_) and a parents’ (*L*_*iP*_) component as

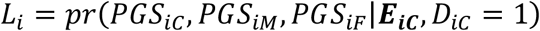

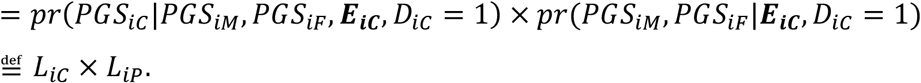

Under the above model, the likelihood components associated with children (*L*_*iC*_) and parental PGS (*L*_*iP*_) data for each family can be derived in terms of the following normal distributions (see Supplemental Notes for detailed derivation):

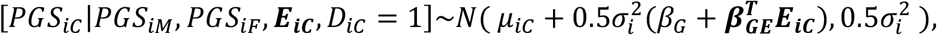

and

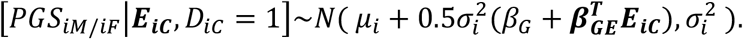

Maximum-likelihood estimation based on 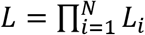 can be complex due to the presence of large dimensional nuisance parameters 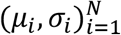. Instead, we propose a combination of likelihood- and moment-based estimation. First, we observe that the likelihood 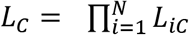 is informative for estimating *β*_*G*_ and ***β***_***GE***_, but one complication is that it requires estimates of family-specific variance parameters 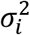. We now observe that under the above model, the PGS values for two parents within each family are expected to have identical distribution, and thus 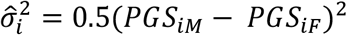 provides an unbiased estimator for 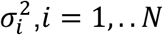. We can now obtain estimates of *β*_*G*_ and ***β***_***GE***_ based on the likelihood *L*_*C*_ with plugged-in values for 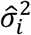. Under the above framework, we show that the final estimator can be derived in an analytic form as a solution to a weighted least-square problem as

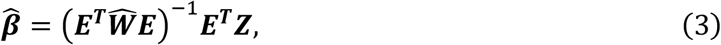

where 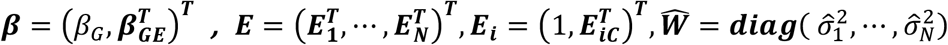, and ***Z*** = 2(*PGS*_1*C*_ − *μ*_1*C*_, …, *PGS*_*NC*_ − *μ*_*NC*_)^*T*^, and *μ*_*iC*_ = 0.5(*PGS*_*iM*_ + *PGS*_*iF*_).

In the special case when no gene-environment interaction terms are incorporated, the estimate of the DE of PGS takes the simple form

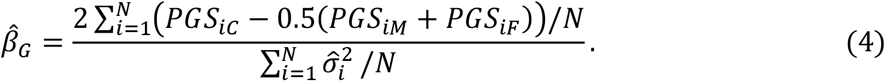

It is noteworthy that the numerator of (4) forms the basis of the pTDT test.^12^ In pTDT, the numerator is normalized by the variance of average PGS values of the parents across families. However, our derivation of (4) suggests that obtaining an unbiased estimate of effect size for PGS in TDT-type analysis requires normalization of the transmission disequilibrium statistics, i.e., the numerator, by an estimate of *within-family* variance.

### Incorporating Indirect Parental Genetic Effects

Next, we extend model (1) to incorporate indirect parental genetic effects as

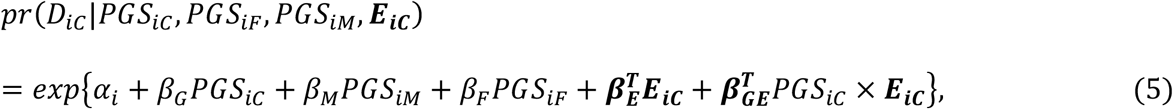

where *β*_*M*_, *β*_*F*_ capture indirect effects of parental PGS on the disease risk of the children not mediated through the children’s genotypes.

The likelihood components associated with children (*L*_*iC*_) remain unchanged after incorporating the indirect genetic effects. We, however, show that the conditional distribution of PGS value in the mother/father in the *i*-th ascertained family, when parental effects are incorporated, needs to be updated as 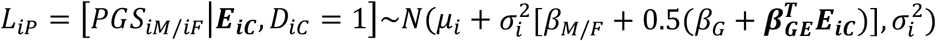. Now we observe that, unlike the previous setting, here the two parents within a family could have asymmetric distribution depending on the difference in the magnitude of their indirect effects (*β*_*M*_ and *β*_*F*_). We can exploit this parental asymmetry in PGS distribution to derive an estimator for the difference of parental indirect genetic effect (*δ*-IDE) as

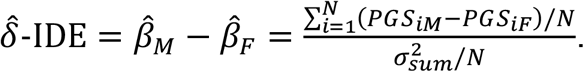

We further show in the presence of indirect effects, an approximately unbiased estimator for 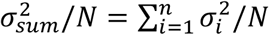 can be derived as

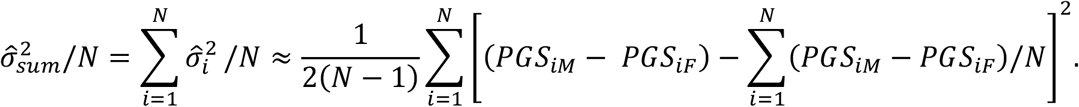

Further, when the indirect effects of parental PGS is incorporated, we observe the form of the estimates of direct effect parameters (***β***) as shown in (3) remains unchanged, but the estimates 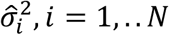 in defining the weight matrix 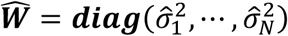 needs to be modified as 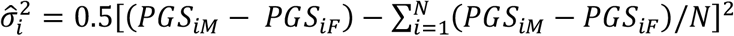 to obtain more accurate estimator of the quantity ***E***^*T*^***WE***.

Detailed derivations of all mathematical results and asymptotic variance estimators are presented in Supplemental Notes.

### Simulation Studies

We conducted 3 types of simulation studies to evaluate the performance of the proposed method under complex population substructure and assortative mating. In the first setting, we simulated PGS values in over 1 million randomly sampled trios from trivariate normal distribution based on Equation (2) and disease status in children under the full model Equation (5), and then further selected families with affected children (*D* = 1). We varied *ρ*_*G*_ = *cor*(*α*_*i*_, *μ*_*i*_) to create different scenarios of population-stratification bias, with a value of 0 indicating no relationship between variation in disease risk and PGS distribution across the underlying substructure – a scenario where we do not expect any bias in population-based association studies. For the investigation of gene-environment interactions, we incorporate a binary (*E*_1_) and a continuous (*E*_2_) variable in the disease model. We allowed complex inter-relationships among baseline disease risk (*α*_*i*_), PGS mean (*μ*_*i*_) and exposure means (*γ*_*i*1_ and *γ*_*i*2_) across families, in manners known to affect the estimation of gene-environment interactions using unrelated individuals. ^15^

We designed the second simulation setting to investigate the robustness of different methods for estimation of simulated effects of educational attainment (EA)-PGS, a score which is known to be highly confounded with environmental factors due to population substructure^16, 17^ in the UK Biobank (UKB) study (www.ukbiobank.ac.uk). We built EA-PGS using independent SNPs (r^2^ < 0.01 within 1000kb) and weights reported in previous work (PGS Catalog ID: PGS002012)^18^ for the UKB participants. To simulate geographical population structure, we matched unrelated UKB males and females within assessment centers (UKB Field ID: 54), and by birth locations defined by north and east coordinates (UKB Field ID: 129 and 130). Additionally, we matched pairs based on both geographical proximity and similarity in educational attainment (UKB Field ID: 6138) to simulate additional effect of assortative mating. For each matched pair of UKB participants, we simulated children by generating their genotype values based on Mendel’s law of transmission and their disease status based on Equation (5), where a “hidden” environmental variable, defined as the average of BMI values (UKB Field ID: 21001) of the two parents, were introduced to influence disease risk.

In the third setting, we used an existing tool *snipar*^19^ to simulate genotype data for 100,000 families across 1,000 independent SNPs, incorporating 20 generations of assortative mating with the parental phenotype correlation at 0.5; and a continuous phenotype influenced by direct genetic effects and assortative mating. To simulate children’s disease status, we used Equation (5), including children’s PGS and parental indirect genetic effects, as well as mid-parental phenotype as a family-level covariate to create an assortative mating effect in children’s disease outcome.

We also conducted additional simulation studies to evaluate potential effects of selection bias on parameter estimates associated with direct and indirect effects obtained from PGS-TRI. Details on all simulation settings, including the matching algorithm used for the UK Biobank simulation, can be found in Supplemental Notes.

### Data Analysis for Application of PGS-TRI to Autism Spectrum Disorders (ASD)

We next illustrated the versatile capabilities of PGS-TRI by using it to analyze genotype and epidemiologic data available on N_trio_ = 18,383 case-parent trios from the Simons Foundation Powering Autism Research for Knowledge (SPARK) study ^13^ (https://sparkforautism.org/) (Table S1). The key objectives included: (1) obtaining transmission-based estimates of the DE of the most advanced ASD-PGS to date built from prior studies, and comparing such effect size from that reported from prior case-control studies, (2) assessing the portability of European-derived PGS to non-European populations across ancestry groups and continuum of genetic ancestry, (3) evaluating the association of PGS for multiple neurocognitive traits with ASD risk using the proposed transmission-based method, (4) characterizing the nature of interaction of ASD-PGS and prenatal exposure on ASD risk, and (5) discovering potential novel association of ASD risk with DE and *δ*-IDE of PGS for gene expression and obesity-related metabolite traits. We allow the ancestry group of each family to be determined by the ancestry group of their offspring (Table S1). Families where parents and the offspring share the same genetic ancestry group are defined as homogeneous families (N_homo_ = 15,876). For most of the analyses presented here, we used homogeneous families; however, for evaluating portability of PGS across genetic ancestry continuum, we included all 18,383 trios.

Specifically, we examined ASD risk associated with direct and indirect effects of pre-constructed PGSs for ASD^20^ and 11 other cognitive and mental health related traits, including educational attainment,^18^ schizophrenia,^21^ strictly defined lifetime major depressive disorder,^22^ bipolar disorder,^23^ bipolar disorder I,^23^ bipolar disorder II,^23^ neuroticism,^18^ insomnia,^18^ chronotype,^18^ body mass index (BMI),^18^ and attention-deficit/hyperactivity disorder (ADHD)^24^ across different ancestry groups. The PGS for ASD itself was defined based on the largest GWAS to date conducted by the iPSYCH consortium and involved a total of 26,637 SNPs. We standardized all the PGSs across the continuum of genetic ancestry by mean adjustment ^25^ of the raw PGSs against the top 5 genetic principal components (PCs), using the 1000 Genomes Project (1kGP) and Human Genome Diversity Project (HGDP) ^26^ as a reference dataset and then divided the PGSs residuals by ancestry-specific standard errors obtained from the reference dataset. This allows interpreting underlying risk parameters, i.e., relative risks, in a standard unit scale across ancestries.

We further examined ASD-PGS by context interactions, defined by the genetic distance on the continuous scale among all populations. Here, genetic distance^27^ is the Euclidean distance of top 5 genetic PCs between each ASD-child and the center of 98 unrelated Finnish individuals from 1kGP+HGDP to represent the PGS training population (North European) from the iPSYCH consortium. Using the homogeneous families, we further examined gene-environment interactions with several prenatal environmental factors, information collected in SPARK participants using questionnaires designed by the Genes and Environment Autism Research Study (GEARS). Finally, we examined the association of ASD risk with genetically predicted levels of gene expression and metabolites using thousands of genetic scores generated by the OMICSPRED project.^28^

Here the underlying hypothesis is that genetically predicted biomarker levels, as captured by the underlying PGS, could affect ASD risk through direct or indirect effects. We do note a caveat that because the PGSs for biomarkers have been derived based on adult samples, direct effect of PGSs on children’s outcome is only possible if the same PGS also predicts biomarker levels in fetal state or/and early childhood. Anticipating limited power for this analysis, we only included those biomolecular traits predicted with accuracy R^2^ ≥ 0.1 by the underlying genetic scores according to the internal validation in OMICSPRED and those included at least 5 SNPs in the underlying model. This criterion resulted in the evaluation of a total of 4,907 gene expression levels and 27 highly correlated obesity-related metabolites. Additional details of data pre-processing and covariate coding can be found in Supplemental Notes.

### Data Analysis for Application of PGS-TRI to Orofacial Clefts (OFCs)

We also applied PGS-TRI to uniquely investigate the polygenic risk of non-syndromic OFCs using case-parent trio data from the Gene Environment Association Studies (GENEVA). ^29, 30^ This analysis included a total of 778 European and 1,126 East-Asian ancestry trios (see Tables S2-3 for distribution of trios by subtypes and exposure). We used these trio data to examine the effects of a pre-defined OFC-PGS on the risk of OFCs across different subtypes and ancestry groups, and its interaction with prenatal exposure to maternal smoking, maternal alcohol consumption, use of multivitamins during pregnancy, and prenatal environmental tobacco smoke exposure. ^29-31^ PGS for cleft lip with or without cleft palate (CL/P) were constructed using 24 SNPs and their respective weights sourced from the PGScatalog.^32, 33^ We standardized the OFC-PGS by ancestry-specific standard errors obtained from the 1000 Genomes Phase 3 Project.^26^ Finally, we also examined the risk of OFCs (CL/P) associated with the DE and *δ*-IDE of genetically predicted gene expressions and metabolite levels using genetic scores generated from the OMICSPRED project. ^28^ Additional details of data pre-processing and covariate coding can be found in Supplemental Notes.

## Results

### Simulation Studies

In simulation studies, where we directly generate data under the assumed model, we observe PGS-TRI produces unbiased effect-size estimates, well-controlled type-I error rates, and calibrated confidence intervals for all of the different types of parameters across a realistic range of population stratification scenarios (Fig.2, Extended Fig.1). The pTDT, being a transmission-based method, also produces unbiased tests for direct genetic effects (DE) and has identical power to PGS-TRI across different scenarios. Standard logistic regression analysis of unrelated case-control participants produces biased inference for DE in the presence of correlations between PGS mean and disease risks across families. Logistic regression and case-only analysis also produce significant bias for inference on gene-environment interaction parameters in the presence of complex population substructures across which disease risk, exposure distribution, and PGS distribution co-vary. If parental genotype data were available for unrelated case-control participants, then logistic regression model could also be used to estimate the magnitude of differential indirect effects (*δ* -IDE). In the absence of population stratification, both methods are valid for the estimation of DE and *δ*-IDE, but logistic regression is more powerful for detecting DE while PGS-TRI is more powerful for detecting *δ*-IDE (Extended Fig.2). Further, logistic regression can produce biased inference for DE not only in the presence of population stratification but also in the presence of *δ*-IDE when parental data are not available to account for such effects (Extended Fig.3).

**Figure 2.**
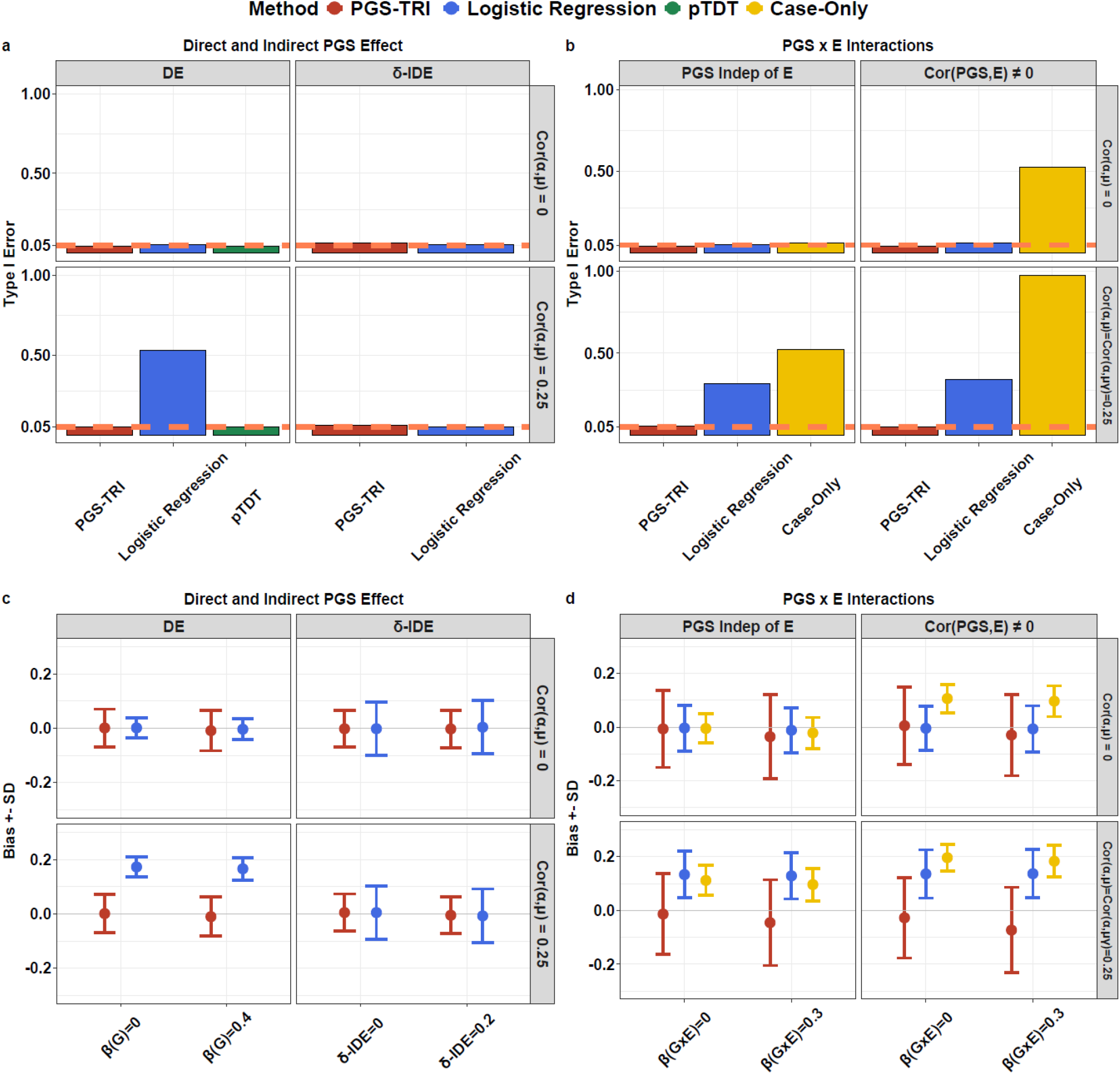
Performance of PGS-TRI and alternative methods for estimating parameters of PGS direct effect (DE), differential parental indirect genetic effect (*δ*-IDE), and PGSxE interactions in simulation studies. Results are shown for (a T pe I error of PGS DE and *δ*-IDE, when underl ing true effects are 0; (b T pe I error of PGS-E interaction, when underl ing true main effect DE is 0.4; (c bias in parameter estimates and magnitude of standard deviation (SD of estimates for DE and *δ*-IDE; (d bias in parameter estimates and magnitude of SD of estimate for PGS-E interaction. pTDT is implemented as an alternative method for testing DE. Logistic regression is implemented for testing and estimation of DE assuming unrelated controls are available of the same size as the number of cases. Logistic regression is also implemented for testing and estimation of *δ*-IDE further assuming parental genot pes are available for the unrelated cases and controls. Further, a case-onl method is also implemented for testing PGSxE interaction. Data are repeatedl simulated for 1000 trios, or 1000 unrelated cases and 1000 unrelated controls from the underl ing population.

We further confirm the robustness of PGS-TRI to realistic patterns of population substructure and assortative mating by considering analyses of educational attainment (EA)-PGS in the UK Biobank study. In this setting, we matched unrelated male and female participants to form “parents” and children’s genotype data were simulated under Mendel’s law of inheritance. We observe when participants are matched by geographical proximity, there is significant across-family correlation (*P* < 5*10^−6^) between EA-PGS and parental-BMI, which is treated as a hidden “environmental” confounder for the simulations of disease risk in children (Extended Fig.4a-4d). In this setting, logistic regression analysis of unrelated cases and controls, even after adjustment for genetic PCs and geographical coordinates, can produce significantly biased inference for DE (Fig.3). When we further added EA to the matching criterion, i.e., allowing for assortative mating in addition to geographical population structure, there was increased bias in logistic regression. PGS-TRI produces both unbiased tests and effect size estimates in all scenarios. Similarly, for the simulation of 20 generations of assortative mating using *snipar*, PGS-TRI remains unbiased for testing and estimation of both PGS direct and indirect effects (Extended Fig. 5).

**Figure 3.**
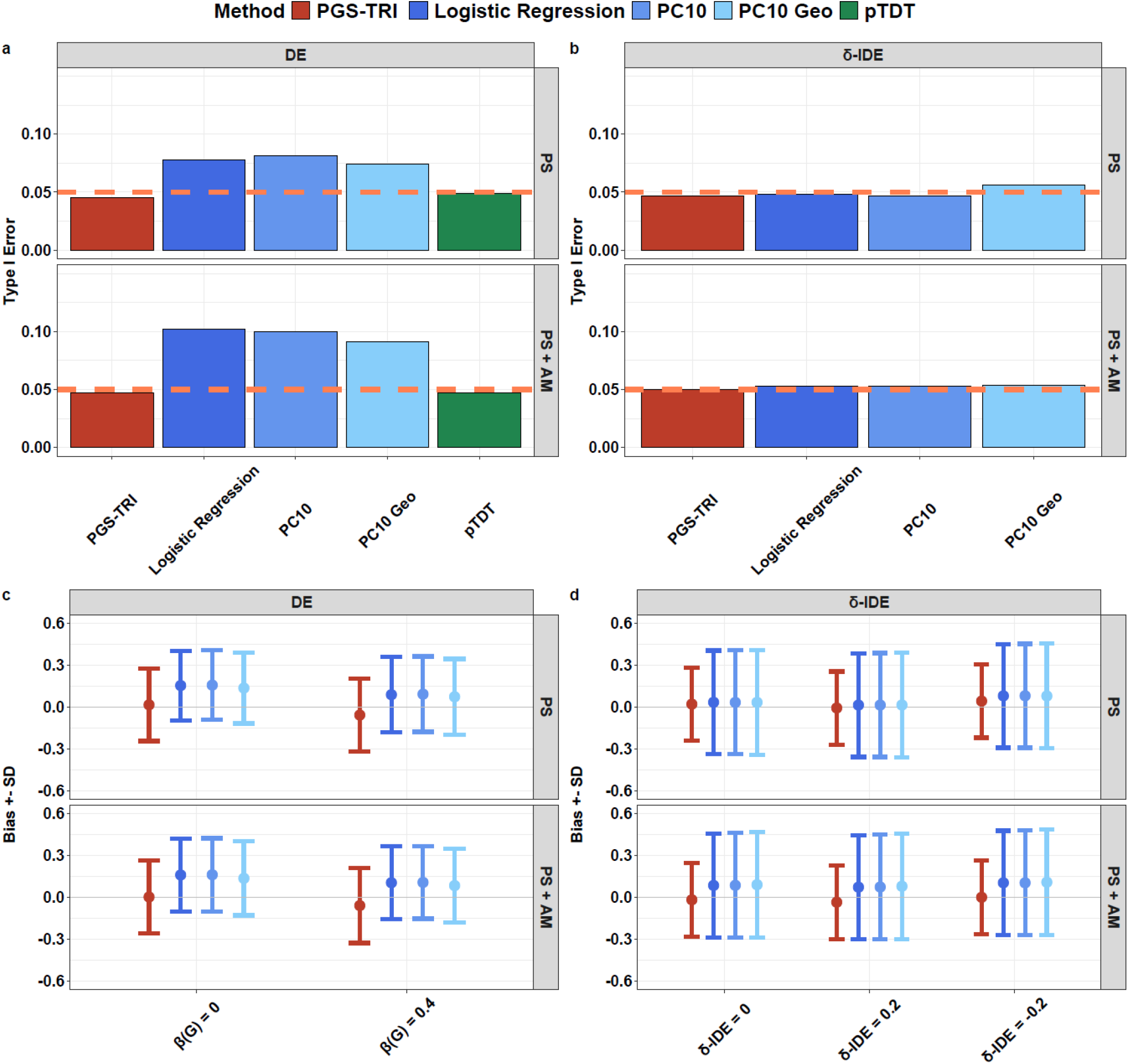
Performance of PGS-TRI and alternative methods for the estimation of genetic effects associated with educational attainment PGS in the UK Biobank-based simulation study. Results are shown for (a T pe I error associated with direct-effect (DE of PGS (b T pe I error associated with differential parental indirect genetic effect (*δ*-IDE of PGS (c bias and SD associated with estimation of DE and (d bias and SD associated with estimation of *δ*-IDE. Among alternative methods, pTDT is implemented for testing of DE. For testing of DE, logistic regression is implemented for the anal sis of unrelated cases and controls without an adjustment, or adjustment for top 10 genetic principal components (PCs constructed from the parental data, or top 10 PCs plus the assessment centres, and north and east birth coordinates of the parents. For the testing and estimation of *δ*-IDE using logistic regression, we assume parental genot pe data are available on unrelated cases and controls. Data on children for matched pairs of UKB participants are repeatedl simulated, and then a set of 2000 case-parent trios, or a set of 2000 unrelated cases and 2000 unrelated controls, are further sampled for subsequent anal sis. Parents were either matched b onl geographic proximit to simulate effect of population stratification or geographical proximit and educational attainment level to simulate effect of both population stratification and assortative mating. **PS** population stratification bias; **AM** assortative mating; **PC10** logistic regression with the top 10 genetic PCs as covariates; **PC10 Geo** logistic regression with the top 10 genetic PCs, north and east birth co-ordinates, and assessment centres as covariates in the model.

### Polygenic Risk for Autism Spectrum Disorders (ASD)

We first examined the association of ASD-PGS derived from the European-ancestry (EUR) iPSYCH study ^20^ with ASD risk across different ancestry groups (Fig.4a; Table S4). We observe for the EUR population (N_trio_ = 12,813), PGS-TRI produced a transmission-based estimate of DE (RR = 1.28, 95% CI = [1.25,1.31]) slightly smaller than the reported estimate (OR = 1.33, 95% CI = [1.30,1.36]) from the iPSYCH study,^20^ which predominantly used unrelated EUR case-control samples. Thus, our estimate of effect size suggests no evidence of significant bias in prior population-based GWAS due to unadjusted population stratification. We also detected evidence of a significant direct effect of the PGS on ASD risk within Americas (N_trio_ = 1,302) and South Asian (N_trio_ = 554) families with the corresponding effect size estimates being of similar magnitude as those derived from EUR families. However, we did not find significant evidence of direct effects of PGS on ASD risk in African (N_trio_ = 792) or East Asian (N_trio_ = 415) families. We did not detect any evidence of IDE associated with ASD-PGS on the risk of ASD.

**Figure 4.**
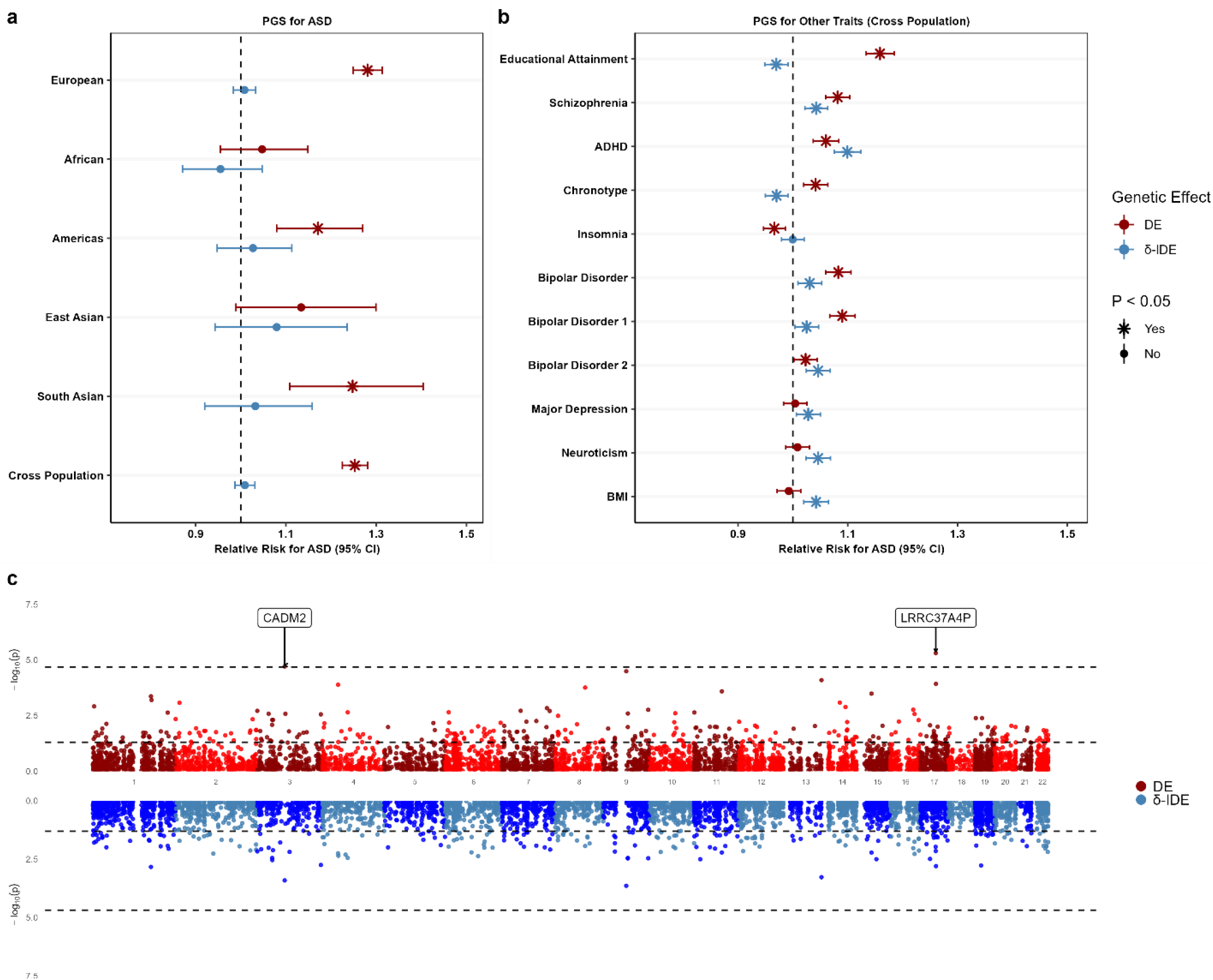
SPARK study results for direct and indirect effects of autism spectrum disorder (ASD). (a Relative risk estimates for direct (DE and maternall medicated indirect effects (*δ*-IDE of ASD-PGS (PGS ID PGS000327 on ASD risk across multiple ancestr groups; (b Relative risk estimates of DE and *δ*-IDE of PGS for multiple neurocognitive traits on ASD risk (onl shown for combined population anal sis due to sample size in ancestral subpopulations ; (c Miami plot showing results from the transcriptome-wide association stud using PGS-TRI of the risk of ASD associated with the DE of PGS for gene-expression traits available from the OMNISPRED stud in homogeneous EUR families. In (a (b, PGS are standardized b subtracting PC projections based on 1000G+HGDP reference data of independent individuals across populations and divided b the population SD calculated using the same reference data so that relative risk (RR corresponds to an increase in risk per SD unit increase in PGS value. **ADHD** attention-deficit/h peractivit disorder; **CI** confidence interval; **BMI** bod mass index, used as a negative control.

Based on the observed pattern of heterogeneity of effect sizes of ASD-PGS across ancestry groups, we hypothesize that the DE of this PGS on ASD risk decreases as the genetic distance between the PGS training population (EUR) and testing sample increases. To validate this, we analyzed data from all families (N_trio_ = 18,383), including those with parents from different ancestries (N_trio_ = 5,570) and examined how of DE of ASD-PGS varies by genetic distance between the analysis sample and training population (Figure 5). The analysis revealed that the log-risk of autism associated with the DE of PGS decreases linearly with increasing genetic distance (Figure 5). This linear relationship is further supported by the highly significant PGS × Context interaction term (*P* < 4*10^−5^). These results remain robust irrespective of whether genetic distances were defined by top 2, 5 or 10 PCs (Table S5).

**Figure 5.**
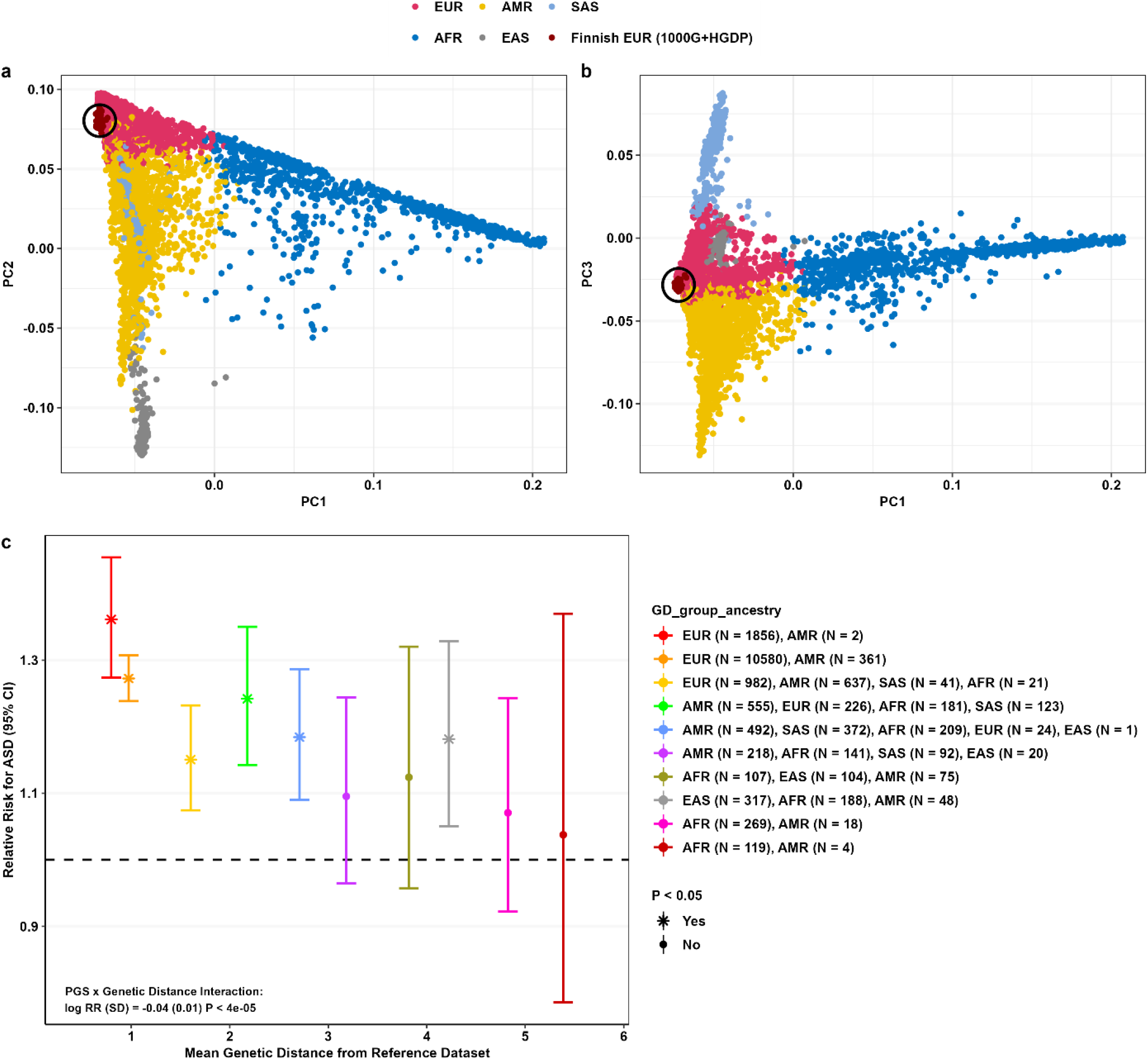
SPARK study results for PGS x context interactions of autism spectrum disorder (ASD). (a (b Principal component (PC anal sis of genetic data in ASD children in SPARK. PCs were projected based on eigenvectors generated using genetics data in 1000G+HGDP unrelated individuals. Dark red points are Finnish individuals from 1000G+HGDP. (c Relative risk estimates of DE for ASD risk using ASD-PGS (PGS ID PGS000327 in 10 groups of equal genetic distance intervals using PGS-TRI. This demonstrates PGS x contect interactions, confirmed b PGS-TRI anal sis (P < 4 * 10^-5. Each group consists of families of different ancestr groups (in total N_trio_ = 18,383. Each ancestr group in the legend notation is based on offspring’s ancestr, note that parents ma be from a different ancestr group. **Genetic distance** is calculated as the Euclidean distance of the top 5 PCs between each ASD child and the center of the unrelated Finnish individuals from 1000G+HGDP reference data. PGS are standardized b subtracting PC projections based on 1000G+HGDP reference data of independent individuals across populations and divided b the population SD calculated using the same reference data so that relative risk (RR corresponds to an increase in risk per SD unit increase in PGS value.

**Figure 6.**
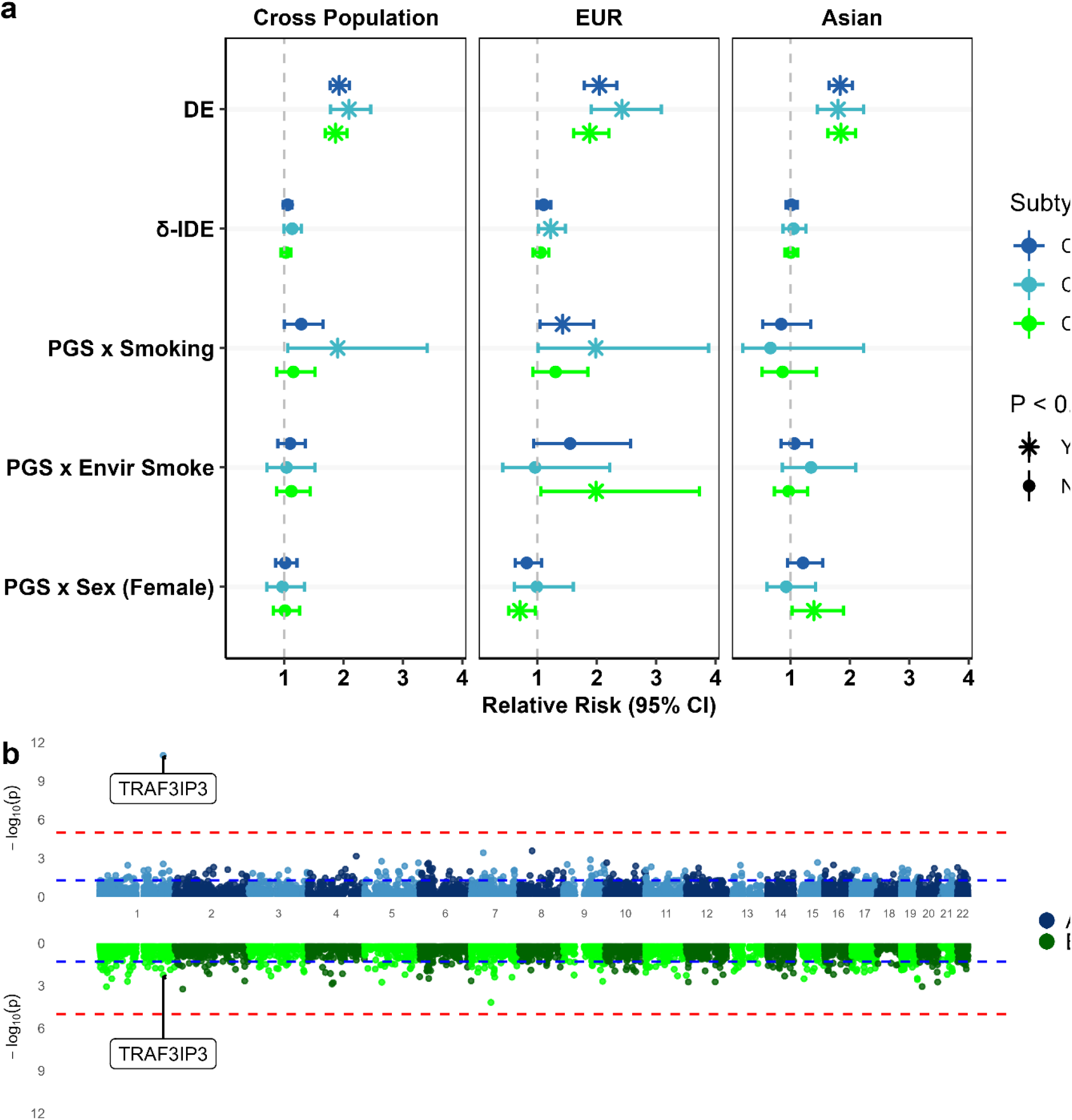
Results from the analysis in the GENEVA study of orofacial clefts (OFCs) trios. (a Estimates of relative risk of different OFC subt pes associated with the DE and *δ*-IDE of OFC-PGS (PGS ID PGS002266 and the interaction of OFC-PGS with several maternall medicated risk factors (b Miami plot showing results from the transcriptome-wide association stud using PGS-TRI of the risk of the combined OFC CL/P associated with the DE of PGS for gene-expression traits available from the OMNISPRED stud. In (a, PGS were standardized within each ancestr group b population mean and SD calculated using 1000G reference data of independent individuals so that relative risk (RR corresponded to an increase in risk per SD unit increase in PGS value.

We explored gene-environment interactions of the ASD-PGS and several specific pre- and peri-natal environmental factors on ASD risk (Table S6). We generally did not observe any strong evidence of interactions, indicating that polygenic risk and environmental factors generally act multiplicatively on the risk of ASD. We observed nominal evidence that maternal alcohol consumption during pregnancy modified the DE of ASD PGS on offspring risk in all families and EUR families (*P* = 0.043 and 0.032, during pregnancy compared to never drinkers).

We next examined the direct effects (DE) and indirect effects (IDE) of PGS for several neurocognitive traits and BMI on ASD risk (Table S7). Results revealed significant DE for most of these scores, with patterns similar to those observed in population-based studies (Figure 4b). As expected, we did not observe any DE of BMI-PGS on autism risk, since children’s genetic predisposition to BMI is not anticipated to influence their own risk for developmental conditions like ASD. Accordingly, BMI serves as a negative control in this analysis. Interestingly, the analysis identified maternally mediated indirect effects (*δ*-IDE > 0) of PGS for multiple psychiatric traits. The corresponding effect sizes suggest that the indirect effects are larger than the direct effects for several traits, including ADHD, bipolar disorder 2, major depression, and neuroticism. We also observed significant evidence of maternally mediated effects of BMI-PGS on ASD risk in children, aligning with epidemiological findings on the influence of maternal obesity on child health outcomes.

Finally, in the analysis of OMICSPRED-generated PGS for gene-expression and metabolite traits, we observed that genetically predicted *CADM2* expression showed significant DE on ASD risk in both cross-ancestry and EUR-only analyses (Figure 4c, Table S8). *CADM2* is predominantly expressed in brain and known to play a crucial role in synapse organization and neuronal activity. Genome-wide association studies have linked common variants near *CADM2* to be associated with a wide variety of psychiatric and neurobehavioral traits,^34-36^ educational attainment,^37^ obesity,^38, 39^ and latent factors underlying insulin resistance and psychiatric traits.^40^ We further validated the results using the precomputed genetic weights^41^ from GTEx v8 in brain tissues and found nominal significance of genetically predicted *CADM2* expression in the substantia nigra (Table S9) on ASD risk. We did not detect evidence (FDR < 0.05) of any indirect effect of either gene-expression or metabolite scores (Table S10-S12). The q-q plots for both DE and IDE associated with 4,907 gene-expression scores show that PGS-TRI controls type-I error rates well in transcriptome-wide analysis (Extended Fig.8) and thus in the future could be used as a discovery tool.

### Polygenic Risk of Orofacial Clefts (OFCs)

We applied PGS-TRI to analyze the risk of orofacial clefts using EUR- and Asian-ancestry trios available from the GENEVA study (Fig.5; Table S13-14). We first examined the risk of various OFCs subtypes with a PGS incorporating 24 SNPs defined by an earlier study (PGS Catalog ID: PGS002266).^32, 33^ We found highly significant and consistent levels of DE of this PGS on the risk of OFCs across different subtypes, including CL-alone, CL&P, and CL/P (CL-alone and CL&P combined), and across both populations. The strength of these associations are of similar magnitude as those reported in previous studies using both population- and family-based samples.^32^ In our analysis, we did not find any evidence of association of this PGS with the CP-alone subtype, an anatomically and embryologically distinct subtype. This was not surprising, considering the original PGS was developed based on studies of CL/P subtypes only. In the cross-ancestry population (combined EUR and Asian population) analysis, we did not observe evidence of differential indirect effects of the parental PGS on the risk OFCs in offspring. In the EUR-only analysis, however, we observed nominal-level evidence of maternally mediated *δ*-IDE of the PGS on the risk of CL-alone (*P* = 0.03).

We further used PGS-TRI to explore the interaction of the DE of OFC-PGS with several known prenatal environmental risk factors and offspring’s sex on the OFCs subtypes (Fig.5a). In the EUR population, we detected evidence of interaction of the PGS with maternal smoking during pregnancy on the risk of CL-alone and combined CL/P; and to a lesser extent with maternal environmental smoking exposure in CL&P. These interactions were not significant in Asians, but the power for detecting interaction in this population was very low as only a small proportion (3%) of the affected probands were exposed to maternal smoking (Table S3). We further observed evidence of PGS by sex interactions in EUR on CL&P risk, but an interaction effect in the opposite direction for the Asian population, which likely cancelled each other in cross-ancestry population analysis.

Finally, we examined the risk of OFCs (CL/P, and combined OFCs) associated with DE and *δ* -IDE of transcriptomic-PGSs generated by the OMICSPRED study.^28^ We detected strong evidence of DE of genetically predicted expression of *TRAF3IP3* on the risk of CL/P (cross-ancestry *P* = 2.0*10^−12^) with the strength of association appearing to be stronger in the Asian population compared with the EUR population (heterogeneity test *P* = 0.01, Fig.5c; Table S15). The genetic score of *TRAF3IP3* expression involved 44 SNPs within the cis-region and had a prediction R^2^ = 0.139 in the EUR population. An intronic SNP rs2235370 of *TRAF3IP3* has been previously reported as a sentinel variant associated with the risk of CL/P in prior GWAS cross-ancestry meta-analysis.^42^ The DE of *TRAF3IP3-*PGS on CL/P risk became insignificant when we removed 8 SNPs in linkage disequilibrium with rs2235370 (r^2^ > 0.3), but the DE of genetic score based only on the 8 SNPs remained highly significant (cross-ancestry *P* = 6.5*10^−13^). Application of genotypic TDT ^43^ further showed a strong association of each of the individual 8 SNPs with the risk of CL/P (Table S16). These results combined suggested the presence of a haplotype in this region which has a protective effect on OFC risk, likely mediated by the expression level of the gene *TRAF3IP3*. We did not detect any evidence of *δ* -IDE of the transcriptomic-PGS on OFCs risk (Extended Fig.9; Table S17). We also did not find evidence of DE or *δ*-IDE of metabolomic-PGSs on OFCs risk (Table S18-19).

## Discussion

We have developed a new analytic framework, PGS-TRI, for the analysis of polygenic scores in case-parent trio studies. It enables transmission-based estimation of the risk of the outcome in offspring, accounting for the direct effects of inherited PGS and its interaction with environmental factors. Further, the method leverages observed asymmetry in PGS value between parents within families to estimate maternally or paternally mediated indirect effects on the offspring outcomes. We conducted simulation studies across various realistic scenarios, including one involving EA-PGS in the UK Biobank study, to demonstrate the robustness of the proposed method against population structure and assortative mating. Our applications of PGS-TRI to 2 early distinct developmental conditions, ASD and OFCs, provide the first transmission-based estimates of effect sizes for established PGSs and address concerns over biases in previous studies caused by uncorrected population structure, assortative mating, or indirect genetic effects. We also applied PGS-TRI to novel analyses exploring polygenic gene-environment interactions on these two conditions. Finally, we used PGSs for gene expression and metabolite traits to examine any evidence of their direct and indirect effects on the two conditions.

Case-parent trio studies and related analytic methods have a significant history and long-standing utility in the field of genetic epidemiology, especially for developmental health conditions in children. Originally, the transmission disequilibrium test (TDT)^44^ was proposed as an allelic test for linkage and association, and was believed that it would form the basis of future GWAS.^45^ Several other key studies noted that transmission-based testing and risk estimation can be conducted based on marker genotype data without having to assume multiplicative effects of underlying alleles.^46, 47^ Subsequently, a series of methods were proposed for the transmission-based analysis of multi-allelic markers,^48^ complex pedigrees,^49^ gene-environment interactions,^50-52^ indirect (also referred to in prior studies as “nurturing”), ^53, 54^ and imprinting/parent-of-origin-effects,^52, 54^ and a more general class of distribution-free methods for family-based association testing also emerged. ^55-57^ As GWAS required very large sample sizes for detecting small polygenic effects, studies based on unrelated cases and controls became widely popular due to the ease of recruiting. In the post-GWAS era, however, there has been now renewed interest in the use of case-parent trios and other family-based studies for more robust characterizations of risks associated with GWAS-identified genetic effects.^7, 11, 12^ Additionally, there are practical considerations that make it more feasible to collect data from parents in families with young affected children than collecting large samples of unrelated cases and controls. Thus, a family trio design is particularly well suited for modern and increasingly common developmental outcomes such as ASD and OFCs, among others.

The first transmission-based method for PGS analysis in case-parent trios was proposed based on the deviation of observed PGS values of children from its expected value under Mendel’s law of transmission, which is represented by the mid-parental PGS values.^12^ While the method provided a valid test, the underlying statistics do not provide valid effect estimates. Here, we demonstrate unbiased risk estimation requires scaling of transmission-disequilibrium statistics by estimates of within-family variance of the PGS, which we now incorporate into the PGS-TRI method. Further, our method, which is motivated by the likelihood of the trio-data, allows for the modeling of PGS-environment interactions within the transmission-based framework and can incorporate indirect effects of parental PGS on children’s outcomes. In particular, we show that under our framework, an estimate of asymmetric maternal and paternal indirect effects of PGS can be obtained by the average difference in PGS values between the two parents across families, scaled by average within-family variance parameters.

We used data from the SPARK consortium to obtain transmission-based estimates of ASD risk associated with pre-defined PGS of ASD. Our results validate previously reported risks in EUR ancestry populations for ASD-PGS. There is also a critical need to assess the portability of ASD-PGS in non-EUR ancestries. Our analysis of the interaction between ASD-PGS and genetic ancestry continuum shows that the transmission-based estimate of effect size for PGS diminishes as an individual’s ancestry deviates further from the training population. While such continuous relationship between PGS effects and genetic ancestry has been shown in population-based studies,^27^ we show for the first time this phenomenon through family-based studies. Our results further show that PGS and the subset of pre- and peri-natal environmental risk factors examined in this study generally have multiplicative effects on the risk of ASD.

We further observed significant direct effects of PGSs for several other neurocognitive traits, including educational attainment, schizophrenia, ADHD, chronotype, insomnia, and bipolar disorders. We also observed evidence of potential indirect effects of parental PGS for many of these same traits on children’s risk of autism spectrum disorder (ASD). However, our simulation studies indicate that asymmetric distributions of PGS between mothers and fathers in trio designs can also result from differential family participation driven by parental traits, such as maternal educational status. Therefore, we cannot exclude the possibility that some indirect effects reflect parental gender-specific selection bias in the SPARK families related to these traits. Further, we identified gender asymmetry in PGS distribution among UK Biobank participants, suggesting potential selection bias in general population cohorts by these traits. However, the magnitude of asymmetry in the UK Biobank was substantially lower than observed among SPARK parents, and for some traits, the direction of asymmetry was even reversed between the two studies (Table S20).

We also applied our new method to a novel investigation of the polygenic risk of OFCs using data from the GENEVA study. This analysis established transmission-based risk estimates for a pre-defined PGS across different OFCs subtypes in both EUR and Asian-ancestry groups. We identified modest evidence of non-multiplicative interaction of the DE of OFC-PGS and maternal smoking, as well as environmental maternal tobacco smoke exposure during pregnancy. Prior genome-wide SNP-environment interaction studies,^29^ including but not limited to GENEVA samples, have not revealed genome-wide significant findings. Joint tests for genetic associations and gene-environment interactions had indicated modest evidence of gene-by-maternal smoking interactions for rs7541797 near PAX7, but little evidence was found for gene-by-environmental tobacco smoking.^29^ In our PGS-based analysis, on the other hand, we find some evidence of aggregated SNPs by smoking interaction effects across different known loci. In the future, large and diverse studies including experimental model systems are needed to further characterize gene-environment interactions in the etiology of OFCs.

While our study has many major strengths as described above, there are also several limitations. Our framework allows estimation of differences in the indirect genetic effects that two parents can exert on children, but it cannot identify the indirect effects of the father and the mother separately. In our setting, we allow family-specific disease risk and PGS distributional parameters to have arbitrary distributions across families. It is possible to extend our framework for estimating the indirect genetic effects for each parent separately by incorporating stronger parametric assumptions. We have modeled gene-environment interactions only with respect to DE of PGS, and further research is merited to extend the model to allow the possibility of interactions of environmental factors with indirect effect of parental PGS on children’s outcome. Our assumption of gene-environment independence within “homogeneous” families is highly robust to the presence of population structure and assortative mating. However, there could be also direct correlation between PGS and environmental exposures due to pleiotropic effects. As discussed before, estimates of indirect effects can be biased in the presence of differential selection bias associated with parental sex. Furthermore, estimates of indirect effects could also be affected by the presence of children who do not live with one or both parents as they will not experience the indirect effect through corresponding parental “exposure”. Finally, the OMICSPRED study primarily trained PGS for molecular traits using adult samples, meaning any DE on developmental outcomes would depend on the PGS being predictive of early molecular traits in children. While molecular QTLs often have robust effects across life stages,^58^ our DE analysis may lack power if early molecular levels are not as well predicted by PGSs generated by OMICSPRED.

The proposed framework for conducting PGS analysis in case-parent trios opens numerous avenues for further research, including applying analogous analyses in other types of ascertained families such as mother-child dyads, case-parent trios with unaffected siblings, and more complex pedigrees which may be ascertained through multiple probands. Additionally, there is potential to extend this model for joint analysis of multiple possibly correlated PGSs, which could be useful for applications such as multivariable Mendelian randomization analysis.^59^ Case-parent trio designs are widely used for many developmental and early childhood conditions due to the feasibility of collecting parental data and biosamples. Moreover, for many late-onset diseases, studies have collected genetic data on extended families ascertained with specific conditions like breast cancer for understanding risks associated with rare high-penetrant mutations. Our proposed method, along with its future extensions, can facilitate PGS analysis in ascertained families, enabling robust characterization of associated risks, both by a PGS itself and in conjunction with rare mutations and non-genetic risk factors.

## Data Availability

The availability of the GWAS summary statistics we used to calculate polygenic scores are detailed in Supplemental Table 5 and downloaded from the PGS Catalog.^33^ The SPARK for ASD genotype data is publicly available through application to the Simons Foundation on SFARI Base at https://base.sfari.org. The SFARI Base Accession ID is “SFARI_DS631850”. GENEVA datasets are available in dbGaP at. https://www.ncbi.nlm.nih.gov through dbGaP accession number phs000094.v1.p1.

## Code Availability

The software PGS-TRI and the polygenic transmission disequilibrium test are publicly available at https://github.com/ziqiaow/PGS.TRI

A website hosting the tutorials is available at https://ziqiaow.github.io/PGS.TRI/

The data analysis and simulations code in this manuscript are available at https://github.com/ziqiaow/PGS-TRI-Analysis

## Acknowledgments

Drs. Wang and Chatterjee were supported by the National Institutes of Health (NIH) grant 1R01HG010480-01. Dr. Wang was also supported by 1K99HG013674-01. Dr. Chatterjee was also supported by U01CA249866, U01HG011719, and 1U24OD023382-01. The GEARS project and Drs. Volk and Ladd-Acosta were supported by funding from R01ES034554 for this work. Dr. Ray was supported by R35GM150836. Drs. Beaty and Ruczinski were supported by NIDCR R01DE031855.

This research has been conducted using the UK Biobank Resource under Application Number 17731.

We are grateful to all of the families in SPARK, the SPARK clinical sites and SPARK staff. We appreciate obtaining access to phenotypic and genetic data on SFARI Base. Approved researchers can obtain the SPARK population dataset described in this study by applying at https://base.sfari.org. We appreciate obtaining access to recruit participants through SPARK research match on SFARI Base.

Funding support for the study entitled “International Consortium to Identify Genes and Interactions Controlling Oral Clefts” was provided by several previous grants from the National Institute of Dental and Craniofacial Research (NIDCR). Data and samples were drawn from several studies awarded to members of this consortium. Funding to support original data collection, previous genotyping and analysis came from several sources to individual investigators. Funding for individual investigators include: R21-DE-013707 and R01-DE-014581 (Beaty); R37-DE-08559 and P50-DE-016215 (Murray, Marazita) and the Iowa Comprehensive Program to Investigate Craniofacial and Dental Anomalies (Murray); R01-DE-09886 (Marazita), R01-DE-012472 (Marazita), R01-DE-014677 (Marazita), R01-DE-016148 (Marazita), R21-DE-016930 (Marazita); R01-DE-013939 (Scott). Parts of this research were supported in part by the Intramural Research Program of the NIH, National Institute of Environmental Health Sciences (Wilcox, Lie). Additional recruitment was supported by the Smile Train Foundation for recruitment in China (Jabs, Beaty, Shi) and a grant from the Korean government (Jee).

The genome-wide association study, also known the Cleft Consortium, is part of the Gene Environment Association Studies (GENEVA) program of the trans-NIH Genes, Environment and Health Initiative [GEI] supported by U01-DE-018993. Genotyping services were provided by the Center for Inherited Disease Research (CIDR). CIDR is fully funded through a federal contract from the National Institutes of Health (NIH) to The Johns Hopkins University, contract number HHSN268200782096C. Funds for genotyping were provided by the NIDCR through CIDR’s NIH contract. Assistance with genotype cleaning, as well as with general study coordination, was provided by the GENEVA Coordinating Center (U01-HG-004446) and by the National Center for Biotechnology Information (NCBI). We sincerely thank all of the patients and families at each recruitment site for participating in this study, and we gratefully acknowledge the invaluable assistance of clinical, field and laboratory staff who contributed to this effort over the years.

Trans-Omics in Precision Medicine (TOPMed) program imputation panel (version Freeze5) was supported by the National Heart, Lung and Blood Institute (NHLBI); see www.nhlbiwgs.org. TOPMed study investigators contributed data to the reference panel, which can be accessed through the Michigan Imputation Server; see https://imputationserver.sph.umich.edu. The panel was constructed and implemented by the TOPMed Informatics Research Center at the University of Michigan (3R01HL-117626–02S1; contract HHSN268201800002I). The TOPMed Data Coordinating Center (3R01HL-120393–02S1; contract HHSN268201800001I) provided additional data management, sample identity checks, and overall program coordination and support. We gratefully acknowledge the studies and participants who provided biological samples and data for TOPMed.

## Extended Figures

**Extended Figure 1.**
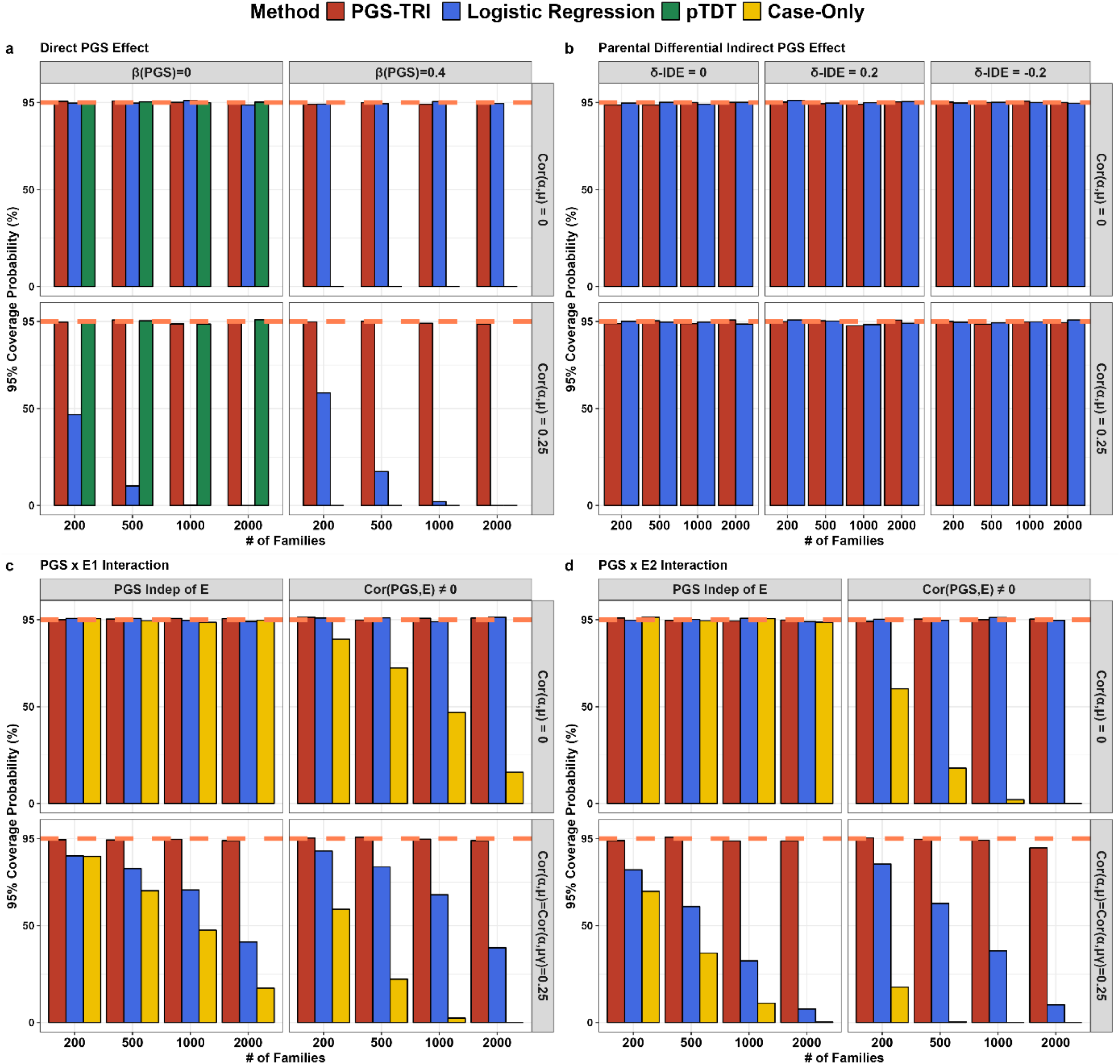
Coverage level of 95% confidence intervals of PGS-TRI and alternative methods for estimation of (a) DE, (b) *δ*-IDE, and (c-d) PGSxE interactions in simulation studies. For each t pe of parameter, results are shown in scenarios in the absence and the presence of population stratification bias. Logistic regression is implemented for the estimation of DE and PGS b E interaction effects assuming unrelated controls are available of the same size as the number of cases. Logistic regression is also implemented for the estimation of *δ*-IDE further assuming that parental genot pes are available for the unrelated cases and controls. Additionall, a case-onl method is implemented for testing PGSxE interaction terms. Data are repeatedl simulated for 1000 trios, or 1000 unrelated cases and 1000 unrelated controls from the underl ing population.

**Extended Figure 2.**
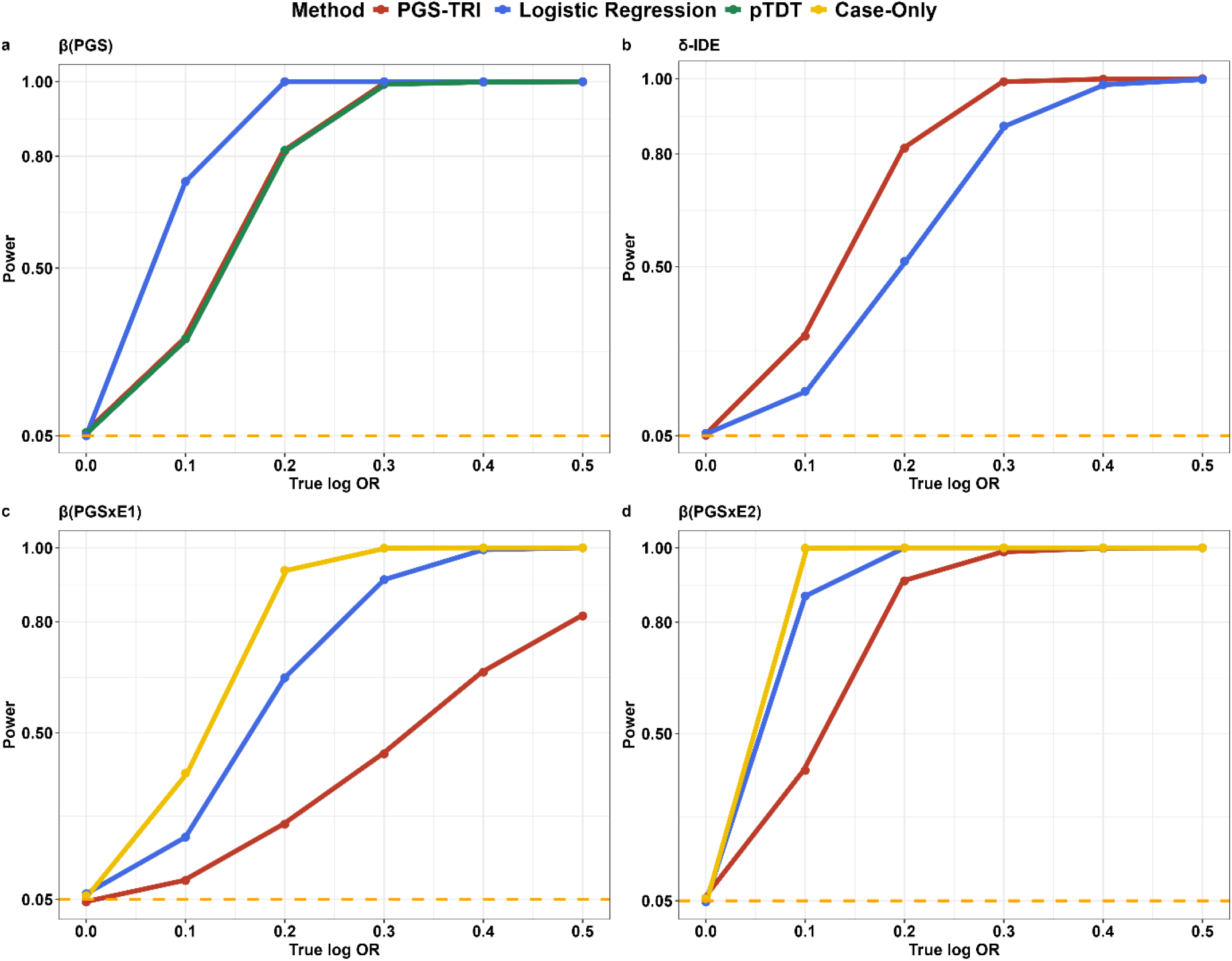
Power of PGS-TRI compared to alternative methods for the detection of (a DE, (b *δ*-IDE, (c (d PGS b E interaction terms. pTDT is implemented as an alternative method for testing DE. The case-onl method is implemented as an alternative method for testing PGSxE. Logistic regression is also implemented for testing of DE and PGSxE assuming unrelated controls are available of the same size as the number of cases. Logistic regression is further implemented for the testing of *δ*-IDE assuming parental genot pes are available for the unrelated cases and controls. For fair comparisons, power results are onl presented in the absence of population stratification when logistic regression and the case-onl method have no bias. Data are repeatedl simulated for 1000 trios, or 1000 unrelated cases and 1000 unrelated controls from the underl ing population. All tests were two-sided and were conducted at a significance level of 0.05.

**Extended Figure 3.**
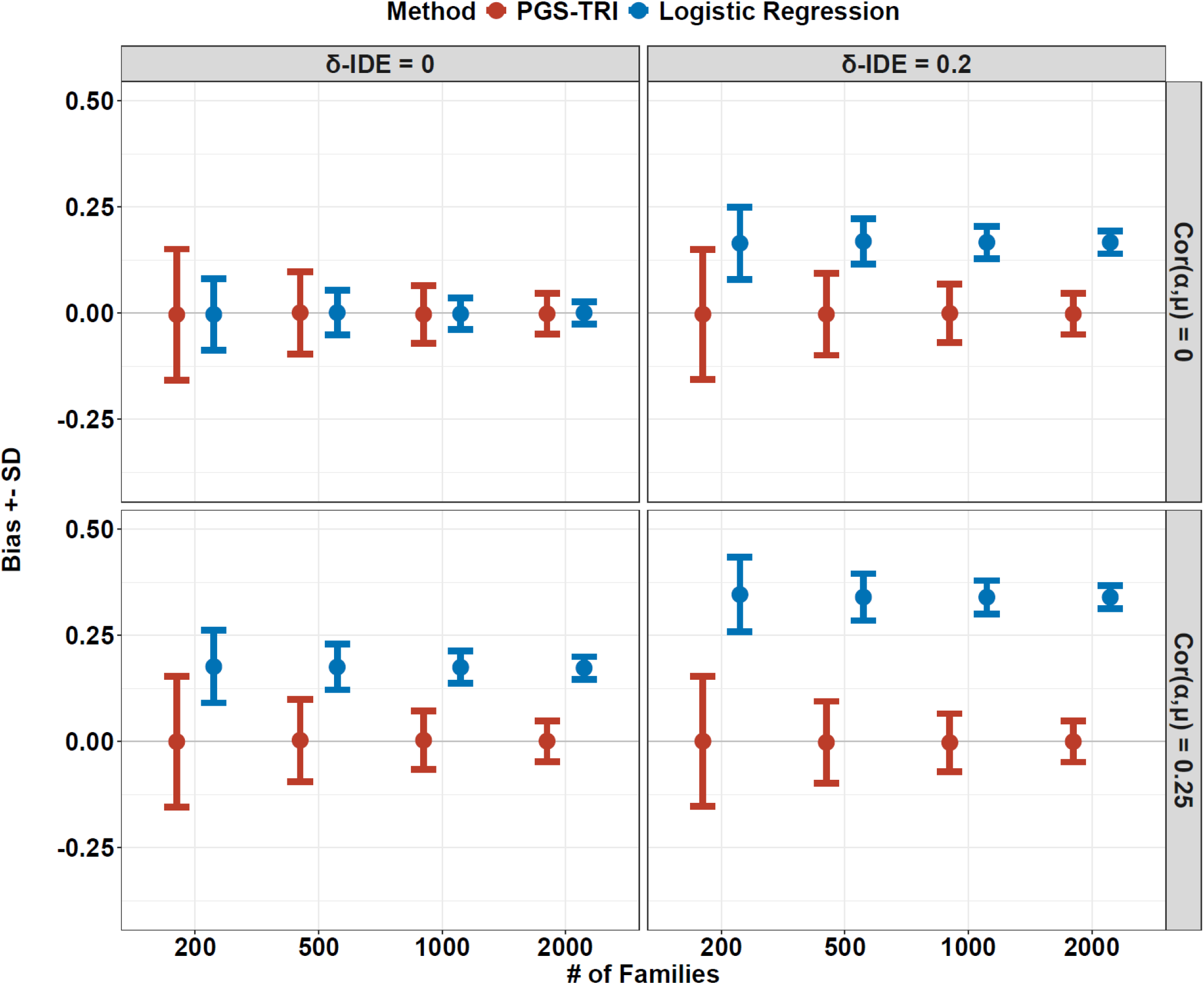
Biases and SD of estimates of DE of PGS using PGS-TRI and logistic regression when *δ*-IDE of parental PGS is not incorporated into modeling. In the top left and bottom left panels, data were simulated assuming no *δ*-IDE. In the top right and bottom right panels, data were simulated in the presence of *δ*-IDE. Both PGS-TRI and logistic regression were fitted without the parental indirect effect parameters in the model.

**Extended Figure 4.**
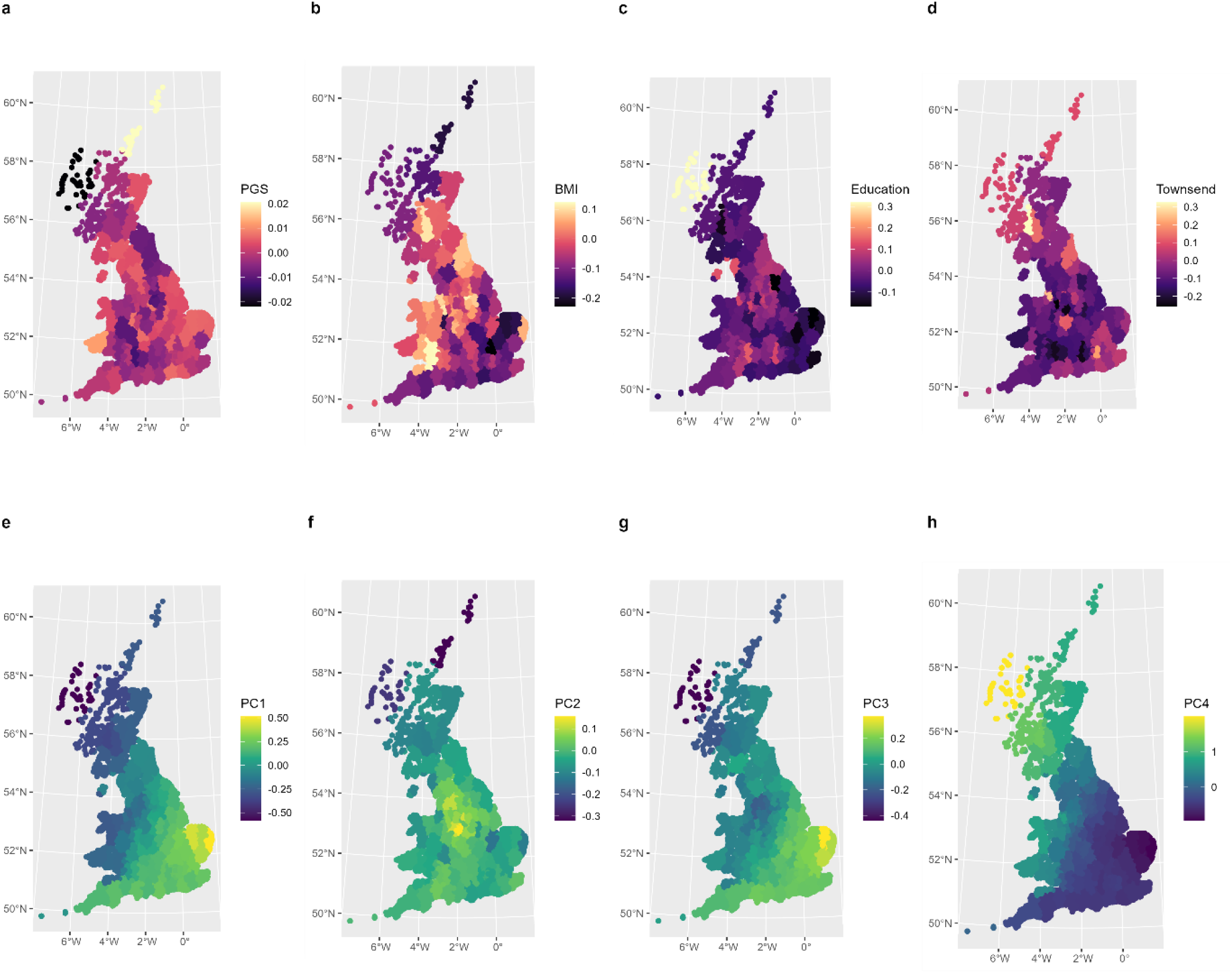
Geographical distributions of (a) Educational Attainment (EA) PGS, (b) Educational Attainment, (c) Body Mass Index (BMI), (d) Townsend Index, (e)-(h)Top 4 Principal Components of unrelated UK Biobank participants (considered as “parents” in our simulation study). Individuals were grouped into 100 clusters based on their east and north co-ordinates of birthplaces using the K-means clustering. Colors represent the mean values of grouped individuals in each cluster on the map. The between-cluster correlation between BMI and EA-PGS is -0.48 (*P* <5*10^−6^, the within-cluster correlation is -0.018 (*P* > 0.05. Parental BMI was treated as a hidden environmental confounding variable while simulating outcome status in children.

**Extended Figure 5.**
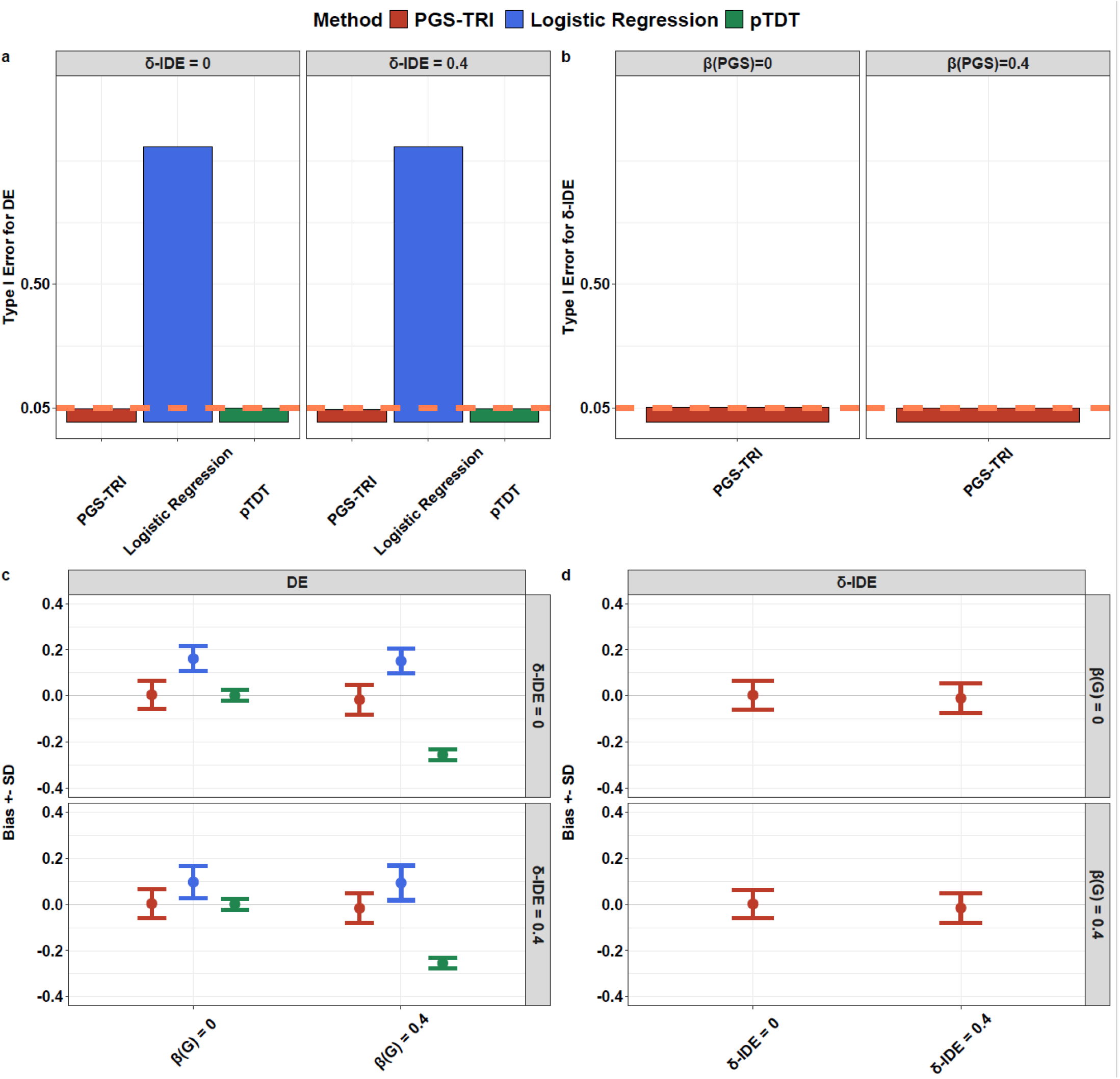
Simulation results under 20 generations of assortative mating effects using *snipar* comparing PGS-TRI, logistic regression, and pTDT. T pe I error for the detection of (a DE when *δ*-IDE = 0 or 0.4, (b *δ*-IDE when DE = 0 or 0.4, biases and SD of estimates of (c DE, (d *δ*-IDE. pTDT is implemented as an alternative method for testing DE. Logistic regression is also implemented for testing of DE assuming unrelated controls are available of the same size as the number of cases. Data are repeatedl simulated for 1000 trios, or 1000 unrelated cases and 1000 unrelated controls from the underl ing population. All tests were two-sided and were conducted at a significance level of 0.05.

**Extended Figure 6.**
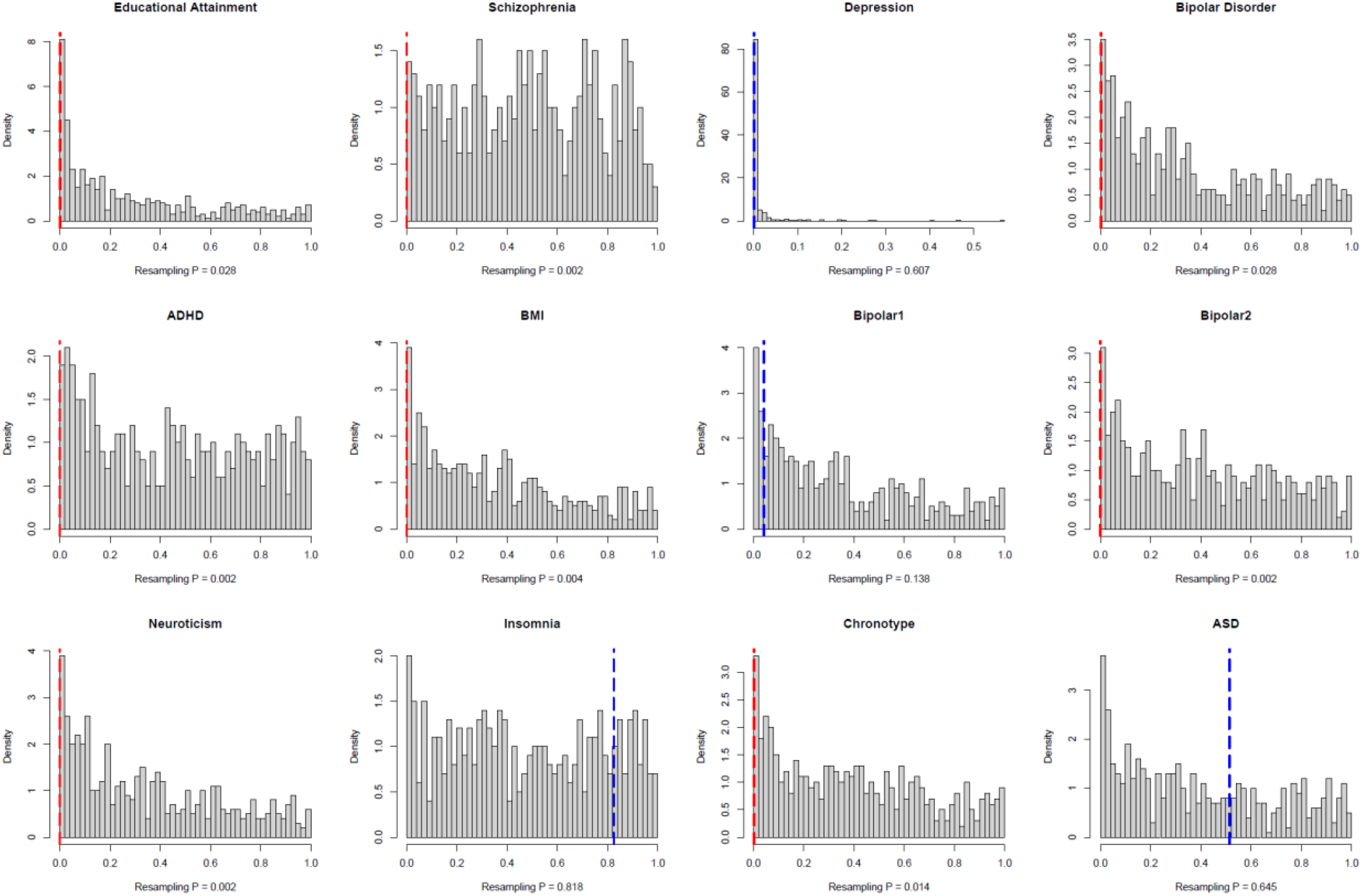
Distributions of 500 resampling P values in comparisons between unrelated European males and females for PGSs of multiple traits in the UK Biobank. Each resampling draws the number of individuals equal to the number of homogeneous European trios in the SPARK consortium (N = 12,813. Dashed line is the P value of *δ*-IDE of each trait from PGS-TRI in SPARK European trios. Red dashed line means significant resampling tests, i.e., the observed *δ*-IDE P value is smaller than 5% of the resampling P.

**Extended Figure 7.**
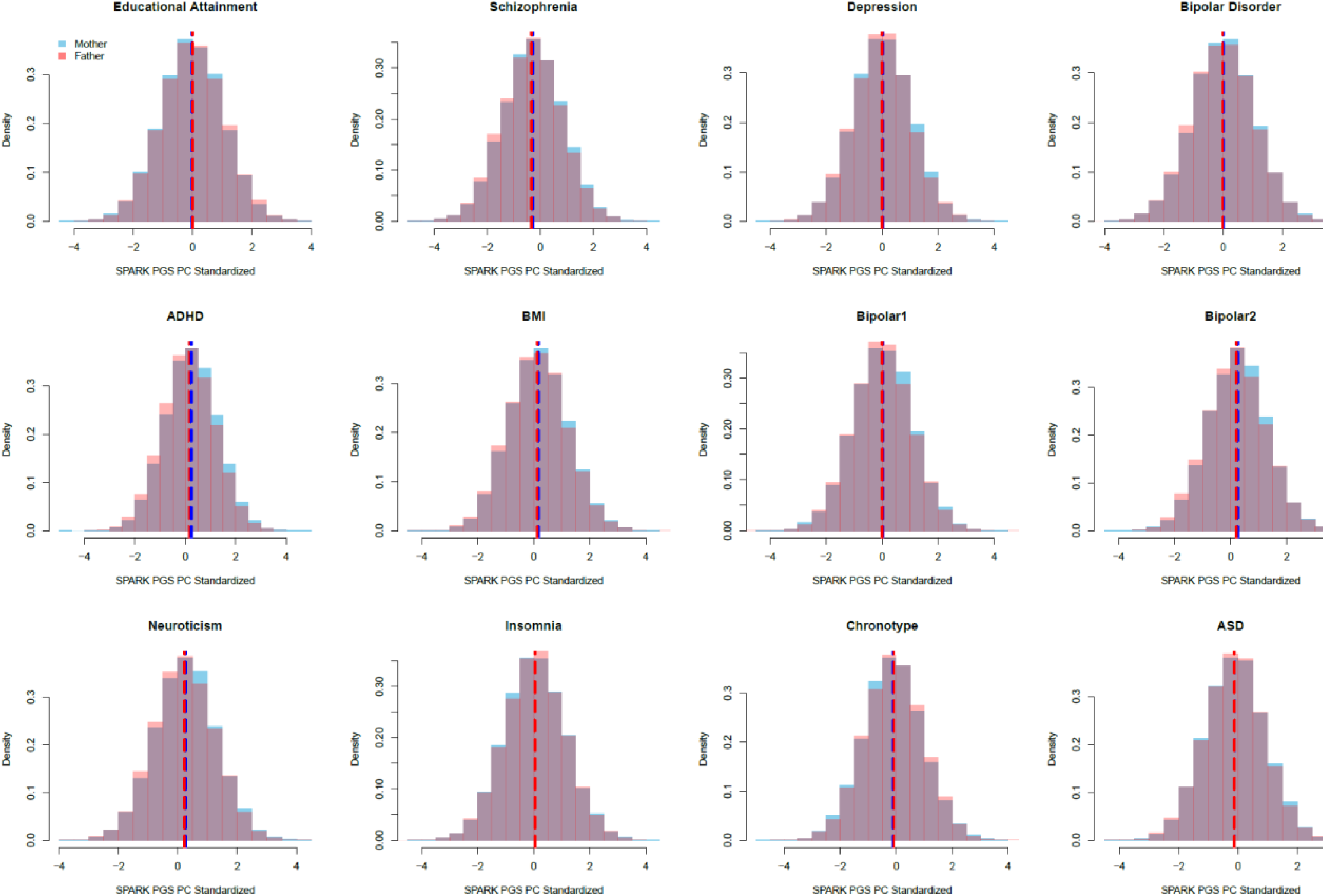
PGS distributions comparisons between mothers and fathers multiple traits in the SPARK consortium among homogeneous European ASD-case parent trios (N_trio_ = 12,813).

**Extended Figure 8.**
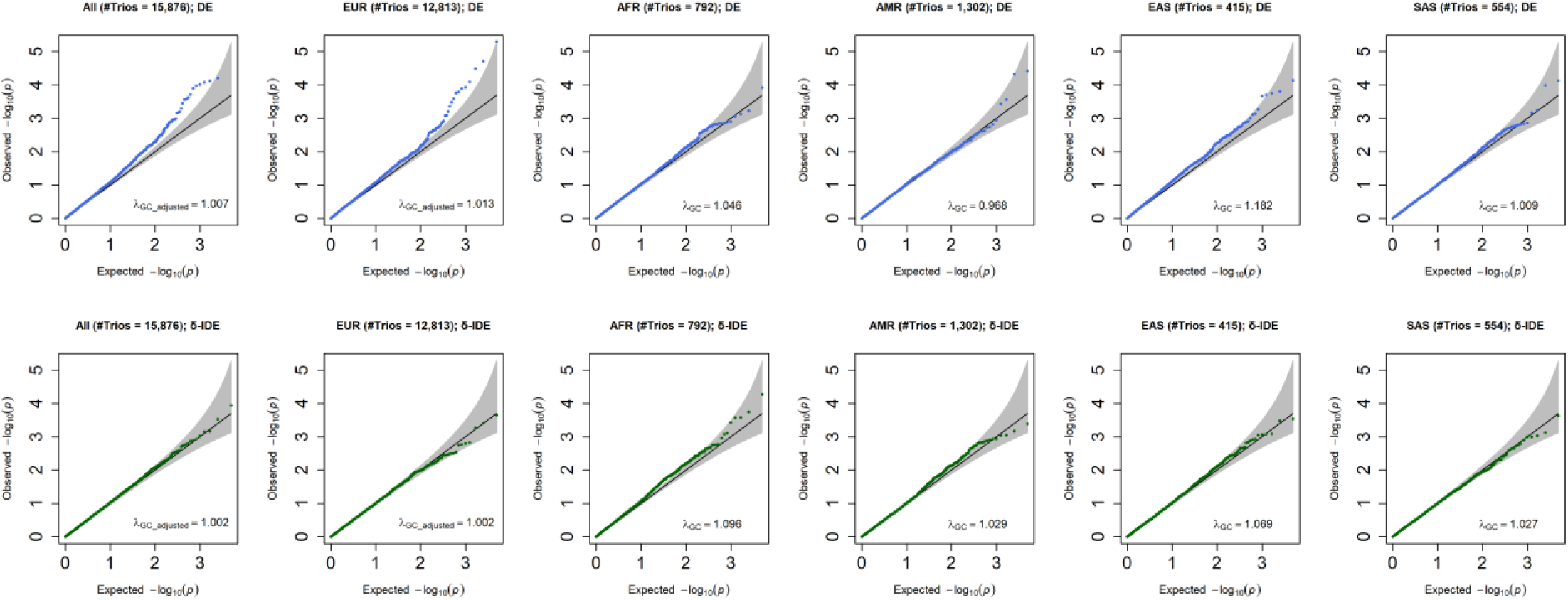
Quantile-quantile plot (QQ plot) of p-values generated by PGS-TRI for the transcriptome-wide association study of autism risk in the SPARK study. The DE and *δ*-IDE of PGS associated with a total of 4,907 transcriptomic traits are tested. The diagonal lines correspond to expected p-values percentiles under the null h pothesis and the shaded regions represent 95% confidence bands. **DE** PGS direct effect; ***δ***-**IDE** differential parental indirect genetic effect; ***λ***_***GC***_**adjusted**_ scaled genomic inflation factor to a stud with 1000 subjects; ***λ***_***GC***_ genomic inflation factor, a value that equals 1 represents no inflation.

**Extended Figure 9.**
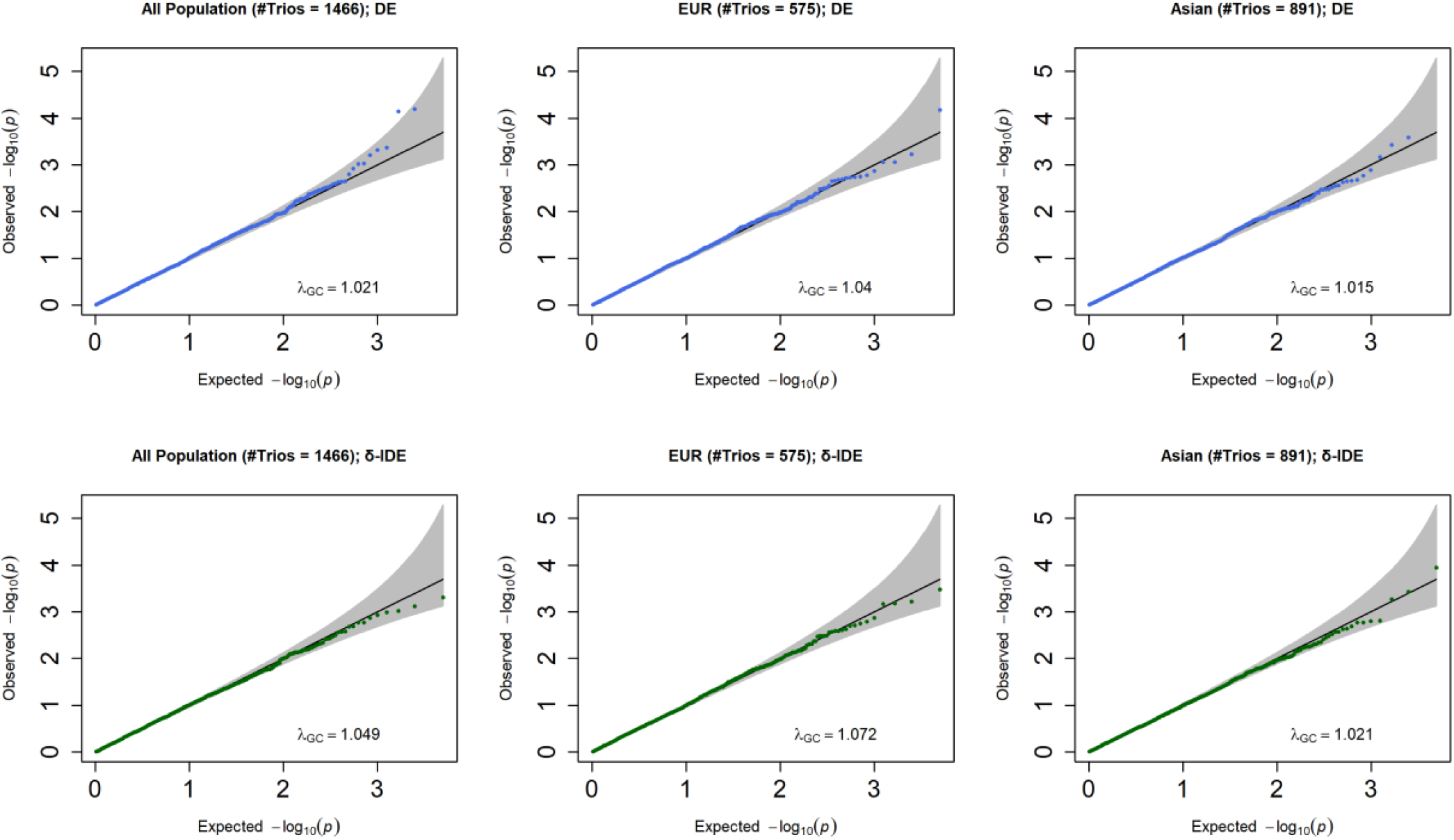
Quantile-quantile plot (QQ plot) of p-values generated by PGS-TRI for the transcriptome-wide association study of the risk of OFC CL/P subtype in the GENEVA study. The DE and *δ*-IDE of PGS associated with a total of 4,991 transcriptomic traits are tested. The diagonal lines correspond to expected p-values percentiles under the null h pothesis and the shaded regions represent 95% confidence bands. **DE** PGS direct effect; ***δ*-IDE** differential parental indirect genetic effect; ***λ***_***GC***_ genomic inflation factor, a value that equals 1 represents no inflation.

