## Supplemental Notes for "Estimation of Direct and Indirect Polygenic Effects and Gene-Environment Interactions using Polygenic Scores in Case-Parent Trio Studies"

### 1 Statistical Derivations

#### 1.1 Derivation under the Assumption of No Indirect Effects of Parental PGS

Let $PGS_{iC}$ denote the PGS value for the child/offspring, $PGS_{iM}$ the PGS for mother, and $PGS_{iF}$ the PGS for father in family $i$, $i = 1, \cdots, N$. Let $D_{iC}$ denote the disease status for the offspring. For each family $i$, the prospective risk model for $D_{iC}$ follows a log-linear model in the form of $\text{pr}(D_{iC}|PGS_{iC}, E_{iC}) = \exp(\alpha_i + \beta_G PGS_{iC} + \boldsymbol{\beta}_{\boldsymbol{E}}^T \boldsymbol{E}_{\boldsymbol{iC}} + \boldsymbol{\beta}_{\boldsymbol{GE}}^T \boldsymbol{E}_{\boldsymbol{iC}} PGS_{iC})$. Under rare disease assumptions, the parameters of the log-linear model can be interpreted as odds ratios, but more generally they correspond to relative risks. Assume that the probability of $(PGS_{iC}, PGS_{iM}, PGS_{iF})^T$ follows

$$\begin{pmatrix} PGS_{iC} \\ PGS_{iM} \\ PGS_{iF} \end{pmatrix} \sim \mathcal{N}\left(\mu_i \mathbb{1}_3, \sigma_i^2 \begin{pmatrix} 1 & \frac{1}{2} & \frac{1}{2} \\ \frac{1}{2} & 1 & 0 \\ \frac{1}{2} & 0 & 1 \end{pmatrix}\right), \tag{1}$$

where $\mu_i$ and $\sigma_i$ are the mean and standard deviation for each family/strata i.

We will show that

$$PGS_{iM/iF}|D_{iC} = 1, \boldsymbol{E}_{\boldsymbol{iC}} \quad \sim \quad \mathcal{N}\left((\mu_i + \frac{\sigma_i^2}{2}(\beta_G + \boldsymbol{\beta}_{\boldsymbol{GE}}^T \boldsymbol{E}_{\boldsymbol{iC}})), \sigma_i^2\right). \tag{2}$$

To derive (2), we consider a risk model of the form

$$\text{pr}(D_{R_j} = 1|U_{R_j}, \boldsymbol{E}_{\boldsymbol{R_j}}) = \exp(\alpha + \beta_G U_{R_j} + \boldsymbol{\beta}_{\boldsymbol{GE}}^T \boldsymbol{E}_{\boldsymbol{R_j}} U_{R_j} + \boldsymbol{\beta}_{\boldsymbol{E}}^T \boldsymbol{E}_{\boldsymbol{R_j}}),$$

where $U_{R_j}$ is the PGS value for offspring and we assume $U_{R_j} \sim \mathcal{N}(\mu, \sigma^2)$. Let $R_i$ and $R_j$ index pair of relatives and $d_{ij}$ denote degree of relatedness. For parent-offspring, $d_{ij} = 1$. We have $cov(U_{R_i}, U_{R_j}) = \rho\sigma^2 = 0.5^{d_{ij}}\sigma^2$. We want to show that

$$U_{R_i}|D_{R_j} = 1, \boldsymbol{E}_{\boldsymbol{R_j}} \quad \sim \quad \mathcal{N}(\mu + 0.5^{d_{ij}}\sigma^2(\beta_G + \boldsymbol{\beta}_{\boldsymbol{GE}}^T \boldsymbol{E}_{\boldsymbol{R_j}}), \sigma^2). \tag{3}$$

We can write the probability as

$$\begin{aligned}
\text{pr}(U_{R_i}|D_{R_j} = 1, \boldsymbol{E}_{\boldsymbol{R_j}}) &= \int_{\mathbb{R}} \text{pr}(U_{R_i}, U_{R_j}|D_{R_j} = 1, \boldsymbol{E}_{\boldsymbol{R_j}}) dU_{R_j} \\
&= \int_{\mathbb{R}} \frac{\text{pr}(D_{R_j} = 1|U_{R_i}, U_{R_j}, \boldsymbol{E}_{\boldsymbol{R_j}}) \text{pr}(U_{R_i}, U_{R_j}|\boldsymbol{E}_{\boldsymbol{R_j}})}{\text{pr}(D_{R_j} = 1|\boldsymbol{E}_{\boldsymbol{R_j}})} dU_{R_j} \\
&= \int_{\mathbb{R}} \frac{\text{pr}(D_{R_j} = 1|U_{R_j}, \boldsymbol{E}_{\boldsymbol{R_j}}) \text{pr}(U_{R_i}, U_{R_j})}{\int_{\mathbb{R}} \text{pr}(D_{R_j} = 1, U_{R_j}|\boldsymbol{E}_{\boldsymbol{R_j}}) dU_{R_j}} dU_{R_j} \\
&= \int_{\mathbb{R}} \frac{\text{pr}(D_{R_j} = 1|U_{R_j}, \boldsymbol{E}_{\boldsymbol{R_j}}) \text{pr}(U_{R_i}, U_{R_j})}{\int_{\mathbb{R}} \text{pr}(D_{R_j} = 1|U_{R_j}, \boldsymbol{E}_{\boldsymbol{R_j}}) f(U_{R_j}) dU_{R_j}} dU_{R_j} \\
&= \int_{\mathbb{R}} \frac{\exp(\alpha + \beta_G U_{R_j} + \boldsymbol{\beta}_{\boldsymbol{GE}}^T \boldsymbol{E}_{\boldsymbol{R_j}} U_{R_j} + \boldsymbol{\beta}_{\boldsymbol{E}}^T \boldsymbol{E}_{\boldsymbol{R_j}}) \text{pr}(U_{R_i}, U_{R_j})}{\int_{\mathbb{R}} \exp(\alpha + \beta_G U_{R_j} + \boldsymbol{\beta}_{\boldsymbol{GE}}^T \boldsymbol{E}_{\boldsymbol{R_j}} U_{R_j} + \boldsymbol{\beta}_{\boldsymbol{E}}^T \boldsymbol{E}_{\boldsymbol{R_j}}) \frac{1}{\sqrt{2\pi\sigma^2}} \exp(-\frac{(U_{R_j} - \mu)^2}{2\sigma^2}) dU_{R_j}} dU_{R_j} \\
&= \int_{\mathbb{R}} \frac{\exp(\beta_G U_{R_j} + \boldsymbol{\beta}_{\boldsymbol{GE}}^T \boldsymbol{E}_{\boldsymbol{R_j}} U_{R_j}) \text{pr}(U_{R_i}, U_{R_j})}{\exp\left(\frac{1}{2}\sigma^2(\beta_G + \boldsymbol{\beta}_{\boldsymbol{GE}}^T \boldsymbol{E}_{\boldsymbol{R_j}})^2 + \mu(\beta_G + \boldsymbol{\beta}_{\boldsymbol{GE}}^T \boldsymbol{E}_{\boldsymbol{R_j}})\right)} dU_{R_j}, 
\end{aligned} \tag{4}$$

we know that $\text{pr}(U_{R_i}, U_{R_j})$ follows a bivariate normal distribution with $cov(U_{R_i}, U_{R_j}) = \rho\sigma^2$:

$$\text{pr}(U_{R_i}, U_{R_j}) = \frac{1}{2\pi\sigma^2\sqrt{1 - \rho^2}} \exp\left\{-\frac{1}{2(1 - \rho^2)}\left[\frac{(U_{R_i} - \mu)^2}{\sigma^2} - 2\rho\frac{(U_{R_i} - \mu)(U_{R_j} - \mu)}{\sigma^2} + \frac{(U_{R_j} - \mu)^2}{\sigma^2}\right]\right\}.$$

For ease of notation, let $x = U_{R_j}$ and $y = U_{R_i}$. To calculate $\int_{\mathbb{R}} \exp(\beta_G x + \beta_{GE} E x) \text{pr}(x, y) dx$, we use conditional distribution of $\text{pr}(X|Y = y)$, i.e.,

$$\begin{aligned}
\int_{\mathbb{R}} \exp(\beta_G x + \beta_{GE} E x) \text{pr}(x, y) dx &= \int_{\mathbb{R}} \exp(\beta_G x + \beta_{GE} E x) \text{pr}(x|Y = y) \text{pr}(y) dx \\
&= \mathbb{E}(e^{(\beta_G + \beta_{GE} E)x}|Y = y) \text{pr}(y)
\end{aligned}$$

We can derive $X|Y = y \sim \mathcal{N}((1-\rho)\mu + \rho y, (1-\rho^2)\sigma^2)$, then we use the moment generating function for normal distribution and we have

$$\int_{\mathbb{R}} \exp(\beta_G x + \beta_{GE} E x) \mathrm{pr}(x, y) dx = \exp\left\{\frac{1}{2}(1-\rho^2)\sigma^2(\beta_G + \beta_{GE}E)^2 + [(1-\rho)\mu + \rho y](\beta_G + \beta_{GE}E)\right\} \frac{1}{\sqrt{2\pi\sigma^2}} \exp\{-\frac{(y-\mu)^2}{2\sigma^2}\}$$
$$= \frac{1}{\sqrt{2\pi\sigma^2}} \exp\left\{-\frac{\{y - [\mu + \rho\sigma^2(\beta_G + \beta_{GE}E)]\}^2}{2\sigma^2}\right\} \exp\left(\mu(\beta_G + \beta_{GE}E) + \frac{\sigma^2(\beta_G + \beta_{GE}E)^2}{2}\right).$$

Plug into (4) and we have proved (3).

From (1), we can derive the conditional probability of $PGS_{iC}$ as

$$PGS_{iC}|PGS_{iM}, PGS_{iF} \sim \mathcal{N}\left(\frac{1}{2}(PGS_{iM} + PGS_{iF}), \frac{\sigma_i^2}{2}\right), \tag{5}$$

denote $\mu_{iC} = \frac{1}{2}(PGS_{iM} + PGS_{iF})$ and $\sigma_{iC}^2 = \frac{\sigma_i^2}{2}$.

##### 1.1.1 Likelihood Derivation

The conditional likelihood for each family $i$ is:

$$\begin{aligned} L_i &= \mathrm{pr}(PGS_{iC}, PGS_{iM}, PGS_{iF}|\boldsymbol{E_{iC}}, D_{iC} = 1) \\ &= L_{iC} \times L_{iP} \\ &= \mathrm{pr}(PGS_{iC}|PGS_{iM}, PGS_{iF}, \boldsymbol{E_{iC}}, D_{iC} = 1) \times \mathrm{pr}(PGS_{iM}, PGS_{iF}|\boldsymbol{E_{iC}}, D_{iC} = 1) \end{aligned}$$

Here,

$$\begin{aligned} L_{iC} &= \mathrm{pr}(PGS_{iC}|PGS_{iM}, PGS_{iF}, \boldsymbol{E_{iC}}, D_{iC} = 1) \\ &= \frac{\mathrm{pr}(D_{iC} = 1|PGS_{iC}, \boldsymbol{E_{iC}})\mathrm{pr}(PGS_{iC}|PGS_{iM}, PGS_{iF})}{\mathrm{pr}(D_{iC} = 1|PGS_{iM}, PGS_{iF}, \boldsymbol{E_{iC}})} \\ &= \frac{\exp(\alpha_i + \beta_G PGS_{iC} + \boldsymbol{\beta_{GE}^T E_{iC}} PGS_{iC} + \boldsymbol{\beta_E^T E_{iC}})\mathrm{pr}(PGS_{iC}|PGS_{iM}, PGS_{iF})}{\int_{-\infty}^{+\infty} \exp(\alpha_i + \beta_G x + \boldsymbol{\beta_{GE}^T} E_{iC} x + \boldsymbol{\beta_E^T E_{iC}}) f_{PGS_C}(x|PGS_{iM}, PGS_{iF}) dx} \\ &= \frac{\exp(\beta_G PGS_{iC} + \boldsymbol{\beta_{GE}^T E_{iC}} PGS_{iC})\mathrm{pr}(PGS_{iC}|PGS_{iM}, PGS_{iF})}{\exp\left\{\mu_{iC}(\beta_G + \boldsymbol{\beta_{GE}^T E_{iC}}) + \frac{1}{2}\sigma_{iC}^2(\beta_G + \boldsymbol{\beta_{GE}^T E_{iC}})^2\right\}} \end{aligned}$$

From (2), we have

$$L_{iP} = \frac{1}{2\pi\sigma_i^2} \exp\left\{-\frac{\left\{PGS_{iM} - [\mu_i + \frac{\sigma_i^2}{2}(\beta_G + \boldsymbol{\beta_{GE}^T E_{iC}})]\right\}^2 + \left\{PGS_{iF} - [\mu_i + \frac{\sigma_i^2}{2}(\beta_G + \boldsymbol{\beta_{GE}^T E_{iC}})]\right\}^2}{2\sigma_i^2}\right\}. \tag{6}$$

##### 1.1.2 Parameter Estimation

We observe that for fixed values of $\beta_G$, $\boldsymbol{\beta_{GE}}$ and $\sigma_i^2, i = 1, \cdots, N$, an unbiased estimator of $\mu_i$ is given by $\hat{\mu_i} = \frac{1}{2}(PGS_{iM} + PGS_{iF}) - \frac{1}{2}\sigma_i^2(\beta_G + \boldsymbol{\beta_{GE} E_{iC}})$, $i = 1, \cdots, N$.

Now by plugging in $\hat{\mu_i}$ to $L_{iP}$, we obtain the profile-likelihood:

$$L_{iP}^* = \frac{1}{2\pi\sigma_i^2} \exp\left\{-\frac{\frac{1}{2}(PGS_{iM} - PGS_{iF})^2}{2\sigma_i^2}\right\}.$$

Therefore the profile-likelihood of each family $i$ is of the form below

$$\begin{aligned} L_i^* &= \mathrm{pr}(PGS_{iC}, PGS_{iM}, PGS_{iF}|\boldsymbol{E_{iC}}, D_{iC} = 1) \\ &= L_{iC} \times L_{iP}^* \\ &= \frac{1}{\sqrt{2\pi\sigma_{iC}^2}} \exp\left\{-\frac{[PGS_{iC} - \mu_{iC} - \sigma_{iC}^2(\beta_G + \boldsymbol{\beta_{GE}^T E_{iC}})]^2}{2\sigma_{iC}^2}\right\} \times \frac{1}{2\pi\sigma_i^2} \exp\left\{-\frac{\frac{1}{2}(PGS_{iM} - PGS_{iF})^2}{2\sigma_i^2}\right\} \end{aligned}$$

The score functions for $\beta_G$ and $\boldsymbol{\beta_{GE}}$ are given by

$$\begin{aligned}
\frac{\partial \log L^*}{\partial \beta_G} &= \sum_{i=1}^{N} \left( PGS_{iC} - \mu_{iC} - \frac{1}{2}\sigma_i^2(\beta_G + \boldsymbol{\beta_{GE}^T E_{iC}}) \right) \\
\frac{\partial \log L^*}{\partial \boldsymbol{\beta_{GE}}} &= \sum_{i=1}^{N} \left( PGS_{iC}\boldsymbol{E_{iC}} - \mu_{iC}\boldsymbol{E_{iC}} - \frac{1}{2}\sigma_i^2(\beta_G + \boldsymbol{\beta_{GE}^T E_{iC}})\boldsymbol{E_{iC}} \right),
\end{aligned}$$

with $\mu_{iC} = \frac{1}{2}(PGS_{iM} + PGS_{iF})$.

We can write the solution in closed form by letting the score functions equal to 0:

$$\sum_{i=1}^{N} \sigma_i^2 \boldsymbol{E_i E_i^T \beta} = 2\sum_{i=1}^{N}(PGS_{iC} - \mu_{iC})\boldsymbol{E_i},$$

where $\boldsymbol{E_i} = (1, \boldsymbol{E_{iC}^T})^T$ and $\boldsymbol{\beta} = (\beta_G, \boldsymbol{\beta_{GE}^T})^T$. Since $\boldsymbol{E_i E_i^T}$ is positive definite (assuming that there is no collinearity between the environmental variables), it is invertible. Therefore, the solution is

$$\hat{\boldsymbol{\beta}} = 2(\sum_{i=1}^{N} \sigma_i^2 \boldsymbol{E_i E_i^T})^{-1} \sum_{i=1}^{N}(PGS_{iC} - \mu_{iC})\boldsymbol{E_i}.$$

In matrix form, this is

$$\hat{\boldsymbol{\beta}} = (\boldsymbol{E}^T \boldsymbol{W} \boldsymbol{E})^{-1} \boldsymbol{E}^T \boldsymbol{Z},$$

where $\boldsymbol{E}_{N\times K} = (\boldsymbol{E_1^T}, \cdots, \boldsymbol{E_N^T})^T, \boldsymbol{W} = \mathbf{diag}(\sigma_1^2, \cdots, \sigma_N^2)$, and $\boldsymbol{Z} = (2(PGS_{1C} - \mu_{1C}), \cdots, 2(PGS_{NC} - \mu_{NC}))^T$.

Finally, we note that in the absence of parental indirect genetic effect, from $L_{iP}$ we can easily show that $E\{\frac{1}{2}(PGS_{iM} - PGS_{iF})^2\} = \sigma_i^2$ and thus throughout we plug in $\hat{\sigma}_i^2 = \frac{1}{2}(PGS_{iM} - PGS_{iF})^2$ for the final estimation.

##### 1.1.3 Asymptotic Variance Estimation when Only Considering Direct PGS Effect

When we only consider the direct PGS effect $\beta_G$, the MLE for $\beta_G$ can be easily derived in closed form by plugging in the method of moments estimator for $\sigma_i^2$:

$$\hat{\beta}_G = \frac{2\sum_{i=1}^{N} \left(PGS_{iC} - \frac{1}{2}(PGS_{iM} + PGS_{iF})\right)/N}{\sum_{i=1}^{N} \hat{\sigma}_i^2/N}.$$

Since we assume that the within family variance $\sigma_i^2$ is finite for $i = 1, \cdots, N$, then it satisfies that

$$\max_{i=1,\cdots,N} \frac{\sigma_i^2}{\sum_{i=1}^{N} \sigma_i^2} \to 0 \quad \text{as} \quad N \to \infty.$$

Therefore by the Lindeberg-Feller central limit theorem, the numerator of the above expression converges in distribution to normal distribution, allowing for the fact that the families are independent but not identically distributed. This holds the same to the denominator since we assume $\sigma_i^4$ is finite. We denote $X := 2\sum_{i=1}^{N} \left(PGS_{iC} - \frac{1}{2}(PGS_{iM} + PGS_{iF})\right)/N$ and $Y := \sum_{i=1}^{N} \frac{1}{2}(PGS_{iM} - PGS_{iF})^2/N$. By definition, the variance of $f(X,Y) = \frac{X}{Y}$ is

$$var(f(X,Y)) = \mathbb{E}\{[f(X,Y) - \mathbb{E}(f(X,Y))]^2\}.$$

The first-order Taylor approximations for $f(X,Y) = \frac{X}{Y}$ around $\boldsymbol{\mu} = (\mu_x, \mu_y) = (\mathbb{E}(X), \mathbb{E}(Y))$ give

$$\mathbb{E}(f(X,Y)) \approx f(\boldsymbol{\mu}),$$

we then have

$$\begin{aligned}
var(f(X,Y)) &\approx \mathbb{E}\left\{[f(X,Y)-f(\boldsymbol{\mu})]^2\right\}\\
&\approx \mathbb{E}\left\{\left[f(\boldsymbol{\mu})+\frac{\partial f(\boldsymbol{\mu})}{\partial x}(X-\mu_x)+\frac{\partial f(\boldsymbol{\mu})}{\partial y}(Y-\mu_y)-f(\boldsymbol{\mu})\right]^2\right\}\\
&= \frac{1}{\mu_y^2}var(X)+\frac{\mu_x^2}{\mu_y^4}var(Y)-\frac{2\mu_x}{\mu_y^3}cov(X,Y)
\end{aligned}$$

Since we have $\mu_x = \beta_G \sum_{i=1}^N \sigma_i^2/N$ and $\mu_y = \sum_{i=1}^N \sigma_i^2/N$, $cov(X,Y)=0$ since $X$ is derived from children's probability conditional on parents, and $Y$ is derived from parents' likelihood term. We can further derive $var(X) = 2\sum_{i=1}^N \sigma_i^2/N^2$ and $var(Y) = 2\sum_{i=1}^N \sigma_i^4/N^2$. Therefore, we have $var(\hat{\beta}_G) = \frac{2}{\sum_{i=1}^N \sigma_i^2} + \frac{2\beta_G^2\sum_{i=1}^N \sigma_i^4}{(\sum_{i=1}^N \sigma_i^2)^2}$. For implementations, we plug-in the estimated $\hat{\sigma}_i^2$ and $\hat{\beta}_G$ and the unbiased estimator for $\hat{\sigma}_i^4 = \frac{1}{12}(PGS_{iM} - PGS_{iF})^4$ into $var(\hat{\beta}_G)$.

##### 1.1.4 Asymptotic Variance Estimation in the General Form

The variance $var(\hat{\boldsymbol{\beta}})$ is obtained using Taylor approximations. Let $\boldsymbol{X} := 2\sum_{i=1}^N (PGS_{iC} - \mu_{iC})\boldsymbol{E_i}/N$ and $\boldsymbol{Y} := \sum_{i=1}^N \hat{\sigma}_i^2 \boldsymbol{E_i}\boldsymbol{E_i}^T/N$. Then we have $f(\boldsymbol{X},\boldsymbol{Y}) = \boldsymbol{Y}^{-1}\boldsymbol{X}$. Similarly to section 1.1.3, we have

$$\mathbb{E}(f(\boldsymbol{X},\boldsymbol{Y})) \approx f(\boldsymbol{\mu}),$$

with $\boldsymbol{\mu} = (\boldsymbol{\mu_x}, \mu_{y_{11}}, \cdots, \mu_{y_{KK}}) = (\mathbb{E}(\boldsymbol{X}), \mathbb{E}(\frac{1}{N}\sum_{i=1}^N \hat{\sigma}_i^2 e_{i1}^2), \mathbb{E}(\frac{1}{N}\sum_{i=1}^N \hat{\sigma}_i^2 e_{i1}e_{i2}), \cdots, \mathbb{E}(\frac{1}{N}\sum_{i=1}^N \hat{\sigma}_i^2 e_{iK}^2))$, with $e_{ij}$ being the $j$-th $(j = 1,\cdots,K)$ environmental variable for children in family $i$ $(i = 1,\cdots,N)$. Here, each $y_{hl}, h \le l = 1,\cdots,K$ is an element in the symmetric matrix $\boldsymbol{Y}$, so there are finite number of parameters in $\boldsymbol{Y}$, i.e., $K(1+K)/2$, $K < \infty$.

By definition,

$$\begin{aligned}
cov(f(\boldsymbol{X},\boldsymbol{Y})) &= \mathbb{E}\left\{[f(\boldsymbol{X},\boldsymbol{Y})-\mathbb{E}(f(\boldsymbol{X},\boldsymbol{Y}))]\,[f(\boldsymbol{X},\boldsymbol{Y})-\mathbb{E}(f(\boldsymbol{X},\boldsymbol{Y}))]^T\right\}\\
&\approx \mathbb{E}\left\{[f(\boldsymbol{X},\boldsymbol{Y})-f(\boldsymbol{\mu})]\,[f(\boldsymbol{X},\boldsymbol{Y})-f(\boldsymbol{\mu})]^T\right\}\\
&\approx \mathbb{E}\left\{\left[\frac{\partial f(\boldsymbol{\mu})}{\partial \boldsymbol{X}}(\boldsymbol{X}-\boldsymbol{\mu_x})+\frac{\partial f(\boldsymbol{\mu})}{\partial y_{11}}(y_{11}-\mu_{y_{11}})+\cdots+\frac{\partial f(\boldsymbol{\mu})}{\partial y_{KK}}(y_{KK}-\mu_{y_{KK}})\right]\cdot\right.\\
&\qquad\left.\left[\frac{\partial f(\boldsymbol{\mu})}{\partial \boldsymbol{X}}(\boldsymbol{X}-\boldsymbol{\mu_x})+\frac{\partial f(\boldsymbol{\mu})}{\partial y_{11}}(y_{11}-\mu_{y_{11}})+\cdots+\frac{\partial f(\boldsymbol{\mu})}{\partial y_{KK}}(y_{KK}-\mu_{y_{KK}})\right]^T\right\}\\
&= \frac{\partial f(\boldsymbol{\mu})}{\partial \boldsymbol{X}}cov(\boldsymbol{X})(\frac{\partial f(\boldsymbol{\mu})}{\partial \boldsymbol{X}})^T+\frac{\partial f(\boldsymbol{\mu})}{\partial y_{11}}var(y_{11})(\frac{\partial f(\boldsymbol{\mu})}{\partial y_{11}})^T+\cdots+\frac{\partial f(\boldsymbol{\mu})}{\partial y_{KK}}var(y_{KK})(\frac{\partial f(\boldsymbol{\mu})}{\partial y_{KK}})^T\\
&+\frac{\partial f(\boldsymbol{\mu})}{\partial y_{11}}cov(y_{11},y_{12})(\frac{\partial f(\boldsymbol{\mu})}{\partial y_{12}})^T+\frac{\partial f(\boldsymbol{\mu})}{\partial y_{12}}cov(y_{11},y_{12})(\frac{\partial f(\boldsymbol{\mu})}{\partial y_{11}})^T+\cdots\\
&+\frac{\partial f(\boldsymbol{\mu})}{\partial y_{K-1,K}}cov(y_{K-1,K},y_{KK})(\frac{\partial f(\boldsymbol{\mu})}{\partial y_{KK}})^T+\frac{\partial f(\boldsymbol{\mu})}{\partial y_{KK}}cov(y_{K-1,K},y_{KK})(\frac{\partial f(\boldsymbol{\mu})}{\partial y_{K-1,K}})^T\\
&= \boldsymbol{\mu_y}^{-1}cov(\boldsymbol{X})(\boldsymbol{\mu_y}^{-1})^T+\sum_{i=1}^N \boldsymbol{\mu_y}^{-1}\boldsymbol{E_i}\boldsymbol{E_i}^T\boldsymbol{\mu_y}^{-1}\boldsymbol{\mu_x}var(\hat{\sigma}_i^2)(\boldsymbol{\mu_y}^{-1}\boldsymbol{E_i}\boldsymbol{E_i}^T\boldsymbol{\mu_y}^{-1}\boldsymbol{\mu_x})^T
\end{aligned}$$

Since $\boldsymbol{X}$ and $\boldsymbol{Y}$ are independent, $cov(\boldsymbol{X}, y_{hl}) = 0$. We also have

$$\boldsymbol{\mu_x} = \sum_{i=1}^{N} \sigma_i^2 (\boldsymbol{\beta}^T \boldsymbol{E_i}) \boldsymbol{E_i}/N = \sum_{i=1}^{N} \sigma_i^2 \boldsymbol{E_i} \boldsymbol{E_i}^T \boldsymbol{\beta}/N$$

$$\boldsymbol{\mu_y} = \sum_{i=1}^{N} \sigma_i^2 \boldsymbol{E_i} \boldsymbol{E_i}^T/N$$

$$cov(\boldsymbol{X}) = 4 \sum_{i=1}^{N} \boldsymbol{E_i} var(PGS_{iC} - \mu_{iC}) \boldsymbol{E_i}^T/N^2 = 2 \sum_{i=1}^{N} \boldsymbol{E_i} \boldsymbol{E_i}^T \sigma_i^2/N^2$$

$$var(\hat{\sigma}_i^2) = 2\sigma_i^4$$

$$\hat{\sigma}_i^4 = \frac{1}{3}(\hat{\sigma}_i^2)^2 = \frac{1}{12}(PGS_{iM} - PGS_{iF})^4$$

##### 1.2 Derivation Incorporating Indirect Effects of Parental PGS

Suppose that the disease outcome of the offspring is also affected by indirect parental PGS effects (IDE), the disease risk model takes the form

$$\begin{aligned}
&\mathrm{pr}(D_{iC} = 1 | PGS_{iC}, PGS_{iM}, PGS_{iF}, \boldsymbol{E_{iC}}) \\
&= \exp(\alpha_i + \beta_G PGS_{iC} + \beta_M PGS_{iM} + \beta_F PGS_{iF} + \boldsymbol{\beta_E^T E_{iC}} + \boldsymbol{\beta_{GE}^T E_{iC}} PGS_{iC}).
\end{aligned}$$

Now the likelihood for each family $i$ becomes

$$\begin{aligned}
L_i &= \mathrm{pr}(PGS_{iC}, PGS_{iM}, PGS_{iF} | \boldsymbol{E_{iC}}, D_{iC} = 1) \\
&= L_{iC} \times L_{iP} \\
&= \mathrm{pr}(PGS_{iC} | PGS_{iM}, PGS_{iF}, \boldsymbol{E_{iC}}, D_{iC} = 1) \times \mathrm{pr}(PGS_{iM}, PGS_{iF} | \boldsymbol{E_{iC}}, D_{iC} = 1) \\
&= \frac{\mathrm{pr}(D_{iC} = 1 | PGS_{iC}, PGS_{iM}, PGS_{iF}, \boldsymbol{E_{iC}}) \mathrm{pr}(PGS_{iC} | PGS_{iM}, PGS_{iF})}{\mathrm{pr}(D_{iC} = 1 | PGS_{iM}, PGS_{iF}, \boldsymbol{E_{iC}})} \times L_{iP} \\
&= \frac{\exp(\beta_G PGS_{iC} + \boldsymbol{\beta_{GE}^T E_{iC}} PGS_{iC}) \mathrm{pr}(PGS_{iC} | PGS_{iM}, PGS_{iF})}{\exp\left\{\mu_{iC}(\beta_G + \boldsymbol{\beta_{GE}^T E_{iC}}) + \frac{1}{2}\sigma_{iC}^2(\beta_G + \boldsymbol{\beta_{GE}^T E_{iC}})^2\right\}} \times L_{iP},
\end{aligned}$$

with $L_{iC}$ unchanged given that the parental effects are canceled out.

We show the below formula

$$PGS_{iM} | \boldsymbol{E_{iC}}, D_{iC} = 1 \quad \sim \quad N\left(\mu_i + \sigma_i^2\left(\beta_M + \frac{1}{2}(\beta_G + \boldsymbol{\beta_{GE}^T E_{iC}})\right), \sigma_i^2\right), \tag{7}$$

$$PGS_{iF} | \boldsymbol{E_{iC}}, D_{iC} = 1 \quad \sim \quad N\left(\mu_i + \sigma_i^2\left(\beta_F + \frac{1}{2}(\beta_G + \boldsymbol{\beta_{GE}^T E_{iC}})\right), \sigma_i^2\right). \tag{8}$$

We derive formula (7) here: for ease of notation, let's denote $x = PGS_{iC}$, $y = PGS_{iM}$, $z = PGS_{iF}$, $E = \boldsymbol{E_{iC}}$, $D = D_{iC}$. Note that $y$ is independent of $z$ and $f(y, z) = f(y)f(z)$.

$$\begin{aligned}
\mathrm{pr}(y, z | E, D = 1) &= \int_{\mathbb{R}} \mathrm{pr}(x', y, z | E, D = 1) dx' \\
&= \int_{\mathbb{R}} \frac{\mathrm{pr}(D = 1 | x', y, z, E) \mathrm{pr}(x', y, z | E)}{\mathrm{pr}(D = 1 | E)} dx' \\
&= \frac{\int_{\mathbb{R}} \mathrm{pr}(D = 1 | x', y, z, E) \mathrm{pr}(x', y, z | E) dx'}{\int\int\int_{\mathbb{R}} \mathrm{pr}(D = 1 | x'', y', z', E) f(x'', y', z') dx'' dy' dz'} \\
&= \frac{\int_{\mathbb{R}} \exp(\beta_G x' + \beta_{GE} E x' + \beta_M y + \beta_F z) f(x', y, z) dx'}{\int\int\int_{\mathbb{R}} \exp(\beta_G x'' + \beta_{GE} E x'' + \beta_M y' + \beta_F z') f(x'', y', z') dx'' dy' dz'}
\end{aligned}$$

Here,

$$\text{numerator} = \int \exp(\beta_G x' + \beta_{GE} E x' + \beta_M y + \beta_F z) f(x'|y,z) dx' f(y) f(z)$$
$$= \exp\{\frac{1}{4}\sigma_i^2(\beta_G + \beta_{GE}E)^2 + \frac{1}{2}(y+z)(\beta_G + \beta_{GE}E)\} \exp(\beta_M y + \beta_F z) f(y) f(z)$$

and

$$\text{denominator} = \exp\left\{\frac{1}{2}\sigma_i^2\Big(\beta_M + \frac{1}{2}(\beta_G + \beta_{GE}E)\Big)^2 + \mu_i\Big(\beta_M + \frac{1}{2}(\beta_G + \beta_{GE}E)\Big)\right\}$$
$$\times \exp\left\{\frac{1}{2}\sigma_i^2\Big(\beta_F + \frac{1}{2}(\beta_G + \beta_{GE}E)\Big)^2 + \mu_i\Big(\beta_F + \frac{1}{2}(\beta_G + \beta_{GE}E)\Big)\right\}$$
$$\times \exp\{\frac{1}{4}\sigma_i^2(\beta_G + \beta_{GE}E)^2\},$$

Then we have

$$\text{pr}(y,z|E,D=1) = \frac{\exp\{[\frac{1}{2}(\beta_G + \beta_{GE}E) + \beta_M]y\} f(y)}{\exp\left\{\frac{1}{2}\sigma_i^2\Big(\beta_M + \frac{1}{2}(\beta_G + \beta_{GE}E)\Big)^2 + \mu_i\Big(\beta_M + \frac{1}{2}(\beta_G + \beta_{GE}E)\Big)\right\}}$$
$$\times \frac{\exp\{[\frac{1}{2}(\beta_G + \beta_{GE}E) + \beta_F]z\} f(z)}{\exp\left\{\frac{1}{2}\sigma_i^2\Big(\beta_F + \frac{1}{2}(\beta_G + \beta_{GE}E)\Big)^2 + \mu_i\Big(\beta_F + \frac{1}{2}(\beta_G + \beta_{GE}E)\Big)\right\}},$$

by plugging in $f(y)$ and $f(z)$ ($y, z \sim N(\mu_i, \sigma_i^2)$), we can easily show formula (7).

Therefore we have

$$L_{iP} = \frac{1}{2\pi\sigma_i^2}\exp\left\{-\frac{\{PGS_{iM} - [\mu_i + \sigma_i^2\,(\beta_M + \frac{1}{2}(\beta_G + \boldsymbol{\beta_{GE}^T E_{iC}}))]\}^2 + \{PGS_{iF} - [\mu_i + \sigma_i^2\,(\beta_F + \frac{1}{2}(\beta_G + \boldsymbol{\beta_{GE}^T E_{iC}}))]\}^2}{2\sigma_i^2}\right\}.$$

##### 1.2.1 Derivation of Estimates of Parental Indirect Genetic Effects

To estimate $\beta_M$ and $\beta_F$, we try to estimate them using the information in $L_{iP}$ alone. We construct a new random variable among case-parent trios

$$X_i = PGS_{iM} - PGS_{iF}$$

Denote $\delta_{MF} = \beta_M - \beta_F$, we have $X_i \sim N(\delta_{MF}\sigma_i^2, 2\sigma_i^2)$ and $\sum_{i=1}^N X_i \sim N(\delta_{MF}\sum_{i=1}^N \sigma_i^2, 2\sum_{i=1}^N \sigma_i^2)$, and thus we have

$$\mathbb{E}(\overline{X}) = \frac{\delta_{MF}\sum_{i=1}^N \sigma_i^2}{N}.$$

Therefore we have the estimator

$$\hat{\delta}_{MF} = \frac{\sum_{i=1}^N X_i/N}{\sum_{i=1}^N \sigma_i^2/N} = \frac{\overline{X}}{\sum_{i=1}^N \sigma_i^2/N}.$$

##### 1.2.2 Approximate Estimator of the Required Scale Factor

We can further derive the expectation of sample variance of $\tilde{X} = (X_1, X_2, \cdots, X_N)$ using information in $L_{iP}$ alone:

$$\mathbb{E}\left(\frac{1}{N-1}\sum_{i=1}^N (X_i - \overline{X})^2\right) = \frac{1}{N-1}\mathbb{E}\left(\sum_{i=1}^N X_i^2 - \frac{(\sum_{i=1}^N X_i)^2}{N}\right) \quad (9)$$
$$= \frac{1}{N}\sum_{i=1}^N 2\sigma_i^2 + \frac{\delta_{MF}^2}{N-1}\left(\sum_{i=1}^N \sigma_i^4 - \frac{1}{N}(\sum_{i=1}^N \sigma_i^2)^2\right), \quad (10)$$

we observe the second term in the above formula will be close to zero if either $\delta_{MF}$ is small or the variability of $\sigma_i^2$ across families is small, or both. Therefore, we can use $\sum_{i=1}^N (X_i - \overline{X})^2/[2(N-1)]$ as an approximately unbiased estimator of $\sigma^2_{sum}/N = \sum_{i=1}^N \sigma_i^2/N$, and thus we estimate $\delta_{MF}$ as

$$\hat{\delta}_{MF} = \frac{\overline{X}}{\sum_{i=1}^N (X_i - \overline{X})^2/[2(N-1)]}.$$

We observe that in the presence of indirect effects, the maximum likelihood estimate of $\boldsymbol{\beta} = (\beta_G, \boldsymbol{\beta}^T_{\boldsymbol{GE}})^T$ obtained from $L_{iC}$ remain unchanged and takes the form $\hat{\boldsymbol{\beta}} = (\boldsymbol{E}^T\boldsymbol{W}\boldsymbol{E})^{-1}\boldsymbol{E}^T\boldsymbol{Z}$, where $\boldsymbol{W} = \mathbf{diag}(\sigma_1^2, \cdots, \sigma_N^2)$. As $\boldsymbol{W}$ is unknown, we need to consider estimation of the elements of $\boldsymbol{E}^T\boldsymbol{W}\boldsymbol{E}$, which are of the form $\sum_{i=1}^N \sigma_i^2 e_{ik}e_{ik'}$, for $k = 1, \cdots, K; k' = 1, \cdots, K$. We observe that for any set of $v_i, i = 1, \cdots, N$, where $v_i = e_{ik}e_{ik'}$ for some $k$ and $k'$, the expectation of the weighted sample variance is

$$\begin{aligned}\mathbb{E}\left(\frac{1}{N-1}\sum_{i=1}^N (X_i - \overline{X})^2 v_i\right) &= \frac{1}{N-1}\sum_{i=1}^N \mathbb{E}\left(X_i^2 v_i + \overline{X}^2 v_i - 2X_i\overline{X}v_i\right) \\ &= \frac{1}{N-1}(\sum_{i=1}^N 2\sigma_i^2 v_i - \frac{4}{N}\sum_{i=1}^N \sigma_i^2 v_i + \frac{2}{N^2}\sum_{i=1}^N \sigma_i^2 \sum_{i=1}^N v_i) + \frac{\delta_{MF}^2}{N-1}\sum_{i=1}^N (\sigma_i^2 - \frac{1}{N}\sum_{i=1}^N \sigma_i^2)^2 v_i\end{aligned}$$

Note that when $v_i = 1$, the above equation is the same as (9). Under the condition $\sum_{i=1}^N v_i = O(N)$,

$$\mathbb{E}\left(\frac{1}{N-1}\sum_{i=1}^N (X_i - \overline{X})^2 v_i\right) \approx \frac{1}{N}\sum_{i=1}^N 2\sigma_i^2 v_i + \frac{\delta_{MF}^2}{N-1}\sum_{i=1}^N (\sigma_i^2 - \frac{1}{N}\sum_{i=1}^N \sigma_i^2)^2 v_i.$$

Above, we again note that if $\delta_{MF}$ is small or $\sigma_i^2$ across families are relatively constant, or both, the last term is expected to be negligible. Thus, we propose using $\sum_{i=1}^N (X_i - \overline{X})^2 v_i/[2(N-1)]$ as an approximate unbiased estimator for $\sum_{i=1}^N \sigma_i^2 v_i/N$.

##### 1.2.3 Asymptotic Variance Estimation

Let $A = \overline{X} = \sum_{i=1}^N (PGS_{iM} - PGS_{iF})/N$ and $B = \sum_{i=1}^N (X_i - \overline{X})^2/[2(N-1)]$. The variance of $\hat{\delta}_{MF}$ can be approximated using first-order Taylor expansion for $\delta_{MF} = f(A, B) = A/B$ around $(\delta_{MF}\sum_{i=1}^N \sigma_i^2/N, \sum_{i=1}^N \sigma_i^2/N)$:

$$var(\hat{\delta}_{MF}) = \frac{2}{\sigma^2_{sum}} + \frac{2\delta_{MF}^2 \sum_{i=1}^N \sigma_i^4}{(\sum_{i=1}^N \sigma_i^2)^2}.$$

Note that the numerator $A$ and the denominator $B$ in $\hat{\delta}_{MF}$ are independent to each other (mean and sample variance of normal distribution). For small $\delta_{MF}$, we can further approximate the variance formula as

$$var(\hat{\delta}_{MF}) = \frac{2}{\sigma^2_{sum}}.$$

Based on the above formula, we can obtain variance estimators by plugging in values for $\hat{\sigma}^2_{sum}$, $\hat{\delta}_{MF}$, and $\sum_{i=1}^N \hat{\sigma}_i^4 = \frac{N^2}{12(N-1)^2}\sum_{i=1}^N (X_i - \overline{X})^4$.

#### 1.3 Estimation when there is selection bias

We show below that the direct effect estimate is unbiased under parental selection bias. Let $S_i$ be the indicator of whether the $i$th family participates in the study or not. We assume the likelihood of selection/participation of a family of child with developmental disorders such as ASD may depend on some factor $x_i$, which could be related to parental characteristics and parents' PGSs for the target trait, but not the PGS value of the children themselves. Mathematically, we assume

$$\begin{aligned}\text{pr}(S_i|x_i, PGS_{iC}, PGS_{iM}, PGS_{iF}, D_{iC} = 1) &= \text{pr}(S_i|x_i, PGS_{iM}, PGS_{iF}, D_{iC} = 1) \\ &= \pi_1(x_i, PGS_{iM}, PGS_{iF}),\end{aligned}$$

where $\pi_1$ denotes probability of selection among family of cases. We also implicitly assume that

$$\text{pr}(PGS_{iC}|PGS_{iM}, PGS_{iF}, x_i) = \text{pr}(PGS_{iC}|PGS_{iM}, PGS_{iF}),$$

i.e., transmission within family in the underlying population is not affected by $x_i$.

Under the above assumptions we can show that

$$\begin{aligned} L_{i,select} &= \text{pr}(PGS_{iC}, PGS_{iM}, PGS_{iF}|D_{iC} = 1, S_i = 1, x_i) \\ &= L_{iC,select} \times L_{iP,select} \\ &= \text{pr}(PGS_{iC}|PGS_{iM}, PGS_{iF}, D_{iC} = 1, S_i = 1, x_i) \times \text{pr}(PGS_{iM}, PGS_{iF}|D_{iC} = 1, S_i = 1, x_i). \end{aligned}$$

Here,

$$\begin{aligned} L_{iC,select} &= \text{pr}(PGS_{iC}|PGS_{iM}, PGS_{iF}, D_{iC} = 1, S_i = 1, x_i) \\ &= \frac{\text{pr}(S_i = 1|PGS_{iC}, PGS_{iM}, PGS_{iF}, D_{iC} = 1, x_i)\text{pr}(PGS_{iC}|PGS_{iM}, PGS_{iF}, D_{iC} = 1, x_i)}{\text{pr}(S_i = 1|PGS_{iM}, PGS_{iF}, D_{iC} = 1, x_i)} \\ &= \text{pr}(PGS_{iC}|PGS_{iM}, PGS_{iF}, D_{iC} = 1) \\ &= L_{iC}, \end{aligned}$$

because the direct effect is estimated through $L_{iC}$ only, the estimate remains unchanged.

We however note that under the above selection mechanism, the following equation

$$E(PGS_{iM} - PGS_{iF}|D_{iC} = 1) = (\beta_M - \beta_F)\sigma_i^2,$$

which is the basis of estimation of indirect effect may not hold anymore. For example, suppose probability of selection of family depends only on mother's choice and such trait is related to $PGS_{iM}$ of the target trait. If we assume a functional relationship of the form

$$\text{pr}(S_i = 1|PGS_{iM}, D_{iC} = 1) = \exp(rPGS_{iM}),$$

then it is easy to see

$$PGS_{iM}|D_{iC} = 1, S_i = 1 \sim N(\mu_i + \sigma_i^2(\beta_M + 0.5\beta_G + r), \sigma_i^2).$$

Therefore, we have

$$E(PGS_{iM} - PGS_{iF}|D_{iC} = 1, S_i = 1) = (\beta_M - \beta_F)\sigma_i^2 + r\sigma_i^2.$$

This implies that asymmetries in parental PGS distributions observed in the sample could reflect differential effects of maternal and paternal PGS on family selection, leading to biased estimates of parental indirect effects.

### 2 Details of Simulation Studies

#### 2.1 Simulate PGS Values Based on the Assumed Model

We directly simulated PGS values for 1,300,000 parent-child trios in the population using a multivariate normal distribution as shown in (1) and prospectively simulated disease status in the children based on a logistic risk model with disease prevalence of 1%. We allow each family $i$ to have specific mean $\mu_i$ and variance $\sigma_i^2$. We let $\sigma_i^2$ follow a mixture of 3-component gamma distributions, reflecting fluctuated variances of PGS values in different families, with the parameters

$$\sigma_i^2 \sim 0.6\Gamma(6, 15) + 0.3\Gamma(15, 30) + 0.1\Gamma(60, 150),$$

so that the mean of $\sigma_i^2$ is approximately 0.4 (this value is close to the population variance of 313-SNP PGS related to breast cancer in UK biobank). Furthermore, we simulated family-specific disease-risk parameters by using a model of the form $\alpha_i \sim N(\alpha + \rho_G \mu_i, 1)$. We varied $\rho_G = cor(\alpha_i, \mu_i)$ to create different scenarios of population-stratification bias, with a value of 0 indicating no relationship between variation in disease risk and PGS distribution across underlying substructure, a scenario where one would not expect any effect of underlying population substructure in creating spurious associations between disease risk and PGS at the population level. We selected different numbers of case-parent trios (N=$200, 500, 1000, 2000$) from a random sample of families by restricting to those families where the children were cases ($D_{iC} = 1$) in the population. The number of trios in the simulation study corresponded to the various sample sizes of different populations in the GENEVA study and the SPARK consortium. We compared PGS-TRI with the pTDT test for the performance of the PGS main effect. We also compared the family-based methods with the performance of population-based case-control studies by randomly sampling unrelated disease-free children from the same simulated family-based population. For the parental indirect effect difference, we included different magnitudes of maternal effects and no paternal effect in the underlying disease risk model and evaluated our model's performance.

For the investigation of the performance of the proposed method for the estimation of gene-environment interaction parameters, for each family, we simulated a binary variable $E_1$ and a continuous variable $E_2$ independent of the underlying PGS values for all three family members. We assume a latent continuous variable $S_1$ for binary $E_1$, and allow $S_1$ and $E_2$ to have family-specific mean values $\gamma_{i1}, \gamma_{i2} \sim N(0, 1)$. The mean distribution of family-specific random effect term $\alpha_i \sim N(\alpha + \rho_G \mu_i + \rho_{GE} \mu_i \gamma_{i1} + \rho_{GE} \mu_i \gamma_{i2}, 1)$ indicates the effect of E and PGS differ by population substructures, with an underlying disease risk model incorporating PGS-environment interaction terms. We compared PGS-TRI with the population-based case-only method to assess the performance of the PGS-E interaction terms. We further let$\gamma_{i1}$ and $\gamma_{i2}$ to co-vary systematically with $\mu_i$ following $cor(\mu_i, \gamma_{ij}) \sim uniform(0, 0.5), j = 1, 2$ to allow for potential population-level correlations between PGS and E due to the effect of population stratification and assortative mating.

#### 2.2 Simulation using the UK Biobank Data

To create realistic population substructures, we simulated offspring genotypes conditional on pairs of independent individuals' genotypes of British white ancestry using the UK Biobank (UKB) genotype data. Each pair was matched within the same assessment centre (UKB Field ID: 54), based on the individual's propensity score generated from the place of birth north and east co-ordinates (UKB Field ID: 129 and 130). Specifically, we used the nearest available Mahalanobis metric matching within 0.1 calipers defined by the propensity score. To further assess the model performances under assortative mating of a single trait, we performed a separate set of matching based on educational attainment (EA) (UKB Field ID: 6138) in addition to the geographical regions. It has been previously reported that EA is a common trait in assortative mating and the EA-PGS are heavily confounded by geographical regions. We built EA-PGS using independent SNPs ($R^2 < 0.01$ within 1000kb) and weights reported in previous work (PGS Catalog ID: PGS002012). We prospectively simulated disease status in the 150 253 unrelated children based on a logistic risk model with a disease prevalence of 2%. We let the intercept term $\alpha_i \sim N(\alpha + \rho_G BMI, 1)$, where BMI values are the baseline values of the mothers in each independent simulated family in UKB (Field ID: 21001). We then compared PGS-TRI with the pTDT test, logistic regression of unrelated individuals, logistic regression adjusting for top 10 genetic principal components (PCs), and additionally adjusting for birth locations and assessment centres for the performance of the PGS main effect by selecting different numbers of case-parent trios ($N = 1000, 2000$) from a random sample of families and the same number of unrelated random controls for the comparisons with logistic regression.

We further grouped the parents into 100 clusters based on their east and north co-ordinates of birthplaces using the K-means clustering. We observed significant correlations (cor $= -0.48$) between BMI and EA-PGS between clusters but not within clusters (cor $= -0.018$). This demonstrated population structure and BMI as the hidden confounding variable which affected the random intercept term in disease risk. We reached the same observations as our first simulation results for PGS main effects. We found that the adjustments of PCs and geographical regions in unrelated individuals showed an improvement compared with logistic regression alone. However, there are still residual biases in unrelated logistic regressions after population substructure adjustments, due to assortative mating, non-linear effects from population structures and geographical regions. Further, PGS-TRI had similar efficiency as logistic regression model adjusted for multiple covariates. We demonstrated that PGS-TRI remained the most unbiased method and produced correct type I error rates.

#### 2.3 Simulation using *snipar*

We also simulated data using existing tool *snipar* [1] and generated 100,000 families across 1,000 independent SNPs. We simulated a continuous phenotype influenced by direct genetic effects and assortative mating. We simulated 20 generations of assortative mating and let the parental phenotype correlation to be 0.5. To simulate children's disease status, we used the full model (11) including children's PGS and parental indirect genetic effects, as well as mid-parental phenotype as a family-level covariate to create an assortative mating effect in children's disease outcome. Mathematically, we let $\alpha_i \sim N(\alpha + cor_G * 0.5(PGS_{iM} + PGS_{iF}), 1)$, and

$$\text{logit pr}(D_{iC}|PGS_{iC}, PGS_{iM}, PGS_{iF}) = \alpha_i + \beta_G PGS_{iC} + \beta_M PGS_{iM} + \beta_F PGS_{iF}. \quad (11)$$

Here, we set $\alpha$ so that the disease prevalence is fixed at around 0.01, and varied the values of $cor_G$ to be 0 and 0.25 to further incorporate assortative mating effects.

#### 2.4 Simulation considering Selection Bias

In addition to the theoretical results we derived in Section 1.3, we further simulated PGS values as described in Section 2.1, assuming that the families with diseased ASD offspring are not randomly selected into the study, i.e.,

$$\text{logit pr}(S_i|PGS_{iM}, PGS_{iF}, E_{i,select}, D_{iC} = 1) = \alpha_{select} + \beta_{M,select} PGS_{iM} + \beta_{F,select} PGS_{iF} + \beta_{E,select} E_{i,select}. \quad (12)$$

Here, $S_i$ indicates whether the family participate into the study, $\beta_{M,select}$ and $\beta_{F,select}$ indicates how mother's and father's PGS values affect the probability of participating into the study, $E_{i,select}$ is a family-level environmental factor that may additionally affect the selection into the study, such as social economic status. Note that when $\alpha_{select} = \beta_{M,select} = \beta_{F,select} = \beta_{E,select} = 0$, the probability $\text{pr}(S_i|PGS_{iM}, PGS_{iF}, E_{i,select}) = 0.5$, which means there is no selection bias and the participation is random. We simulated 8 scenarios when

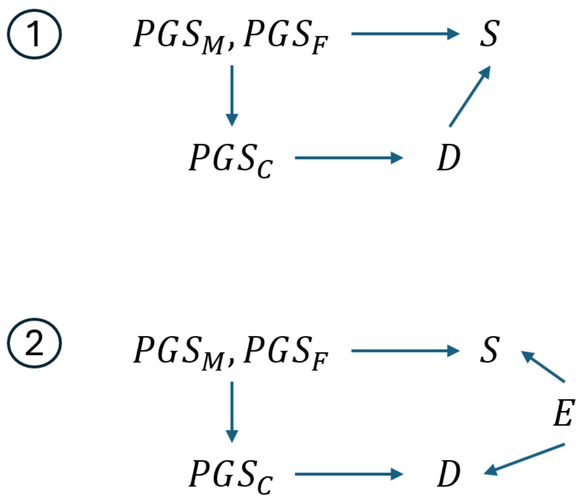

Figure 1: Simulation scenarios for alternative selection bias mechanisms for case-parent trio sampling.

we assumed parents' PGSs can influence the family participation into the study, as shown in Fig 1, corresponding to equation(12). Here $D$ is the children's disease status, and $S$ is selection into the study, $E$ is an environmental factor that has an effect on both $S$ and $D$. For both (1) and (2) in Fig.1, we set equal parental selection effect $\beta_{M,select} = \beta_{F,select} = 0.2$ or different parental selection effect $\beta_{M,select} = 0.2, \beta_{F,select} = 0$.

In (2), we assumed strong environmental factor selection effect $\beta_{E,select} = 0.4$. For the offspring disease risk, we assumed $\beta_G = 0$ or $\beta_G = 0.4$. For all 8 scenarios, we randomly selected 1000 numbers of case-parent trios from families where the children were cases ($D_{iC} = 1$) and then further sub-sample families based on the selection probability mechanism described in equation(12).

We found that when parents' trait-related PGS values have an effect on the participation rate into the study, there is no bias or inflation in the estimation of DE; for $\delta$-IDE, when this selection effects are equal between the two parents, then the estimation remains correct, but when the two parents have different selection effects, then it will cause bias on the estimate of $\delta$-IDE (Fig.2 & 3). Note that this conclusion is consistent with previous research on selection bias in logistic regression [2,3].

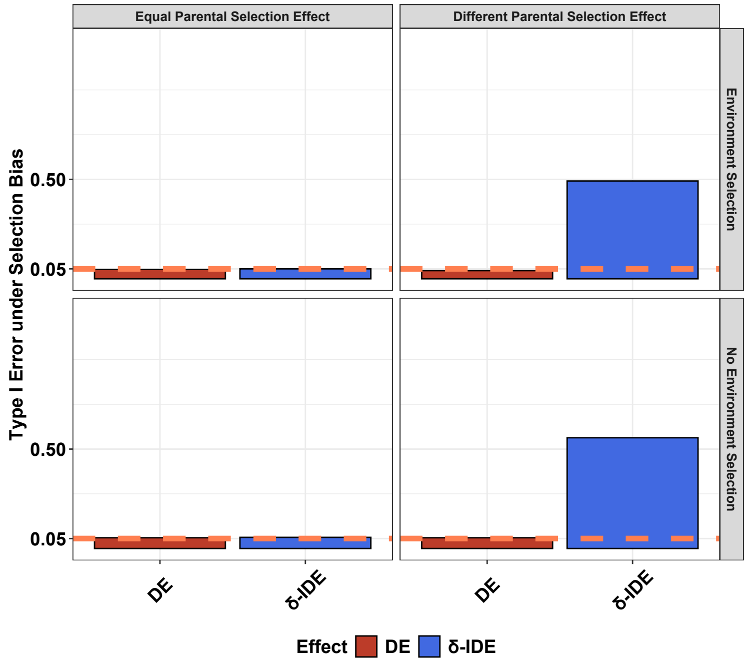

Figure 2: Type I error of the simulation study when there are selection bias in the study of 1000 families. Equal parental selection effect is when $\beta_{M,select} = \beta_{F,select} = 0.2$, different parental selection effect is when $\beta_{M,select} = 0.2, \beta_{F,select} = 0$. Environment selection effect is $\beta_{E,select} = 0.4$.

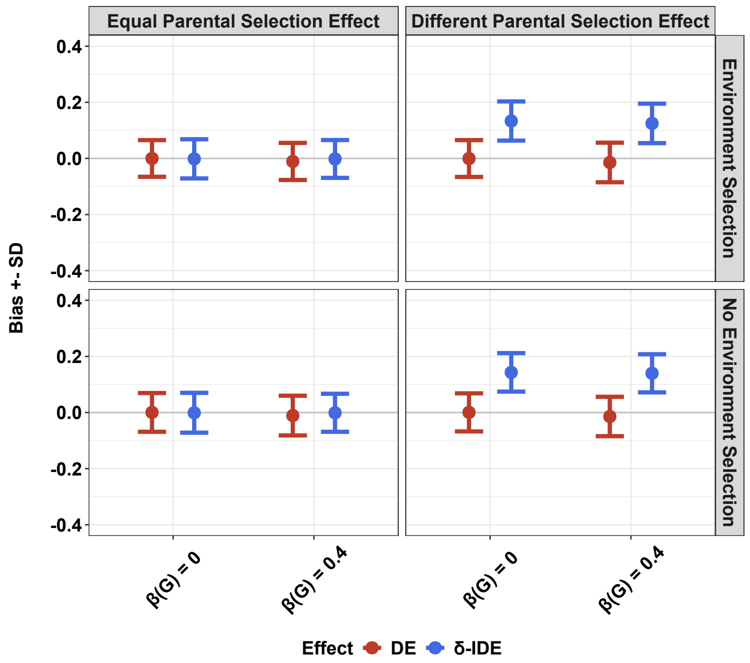

Figure 3: Bias and SD of the simulation study when there are selection bias in the study of 1000 families. Equal parental selection effect is when $\beta_{M,select} = \beta_{F,select} = 0.2$, different parental selection effect is when $\beta_{M,select} = 0.2, \beta_{F,select} = 0$. Environment selection effect is $\beta_{E,select} = 0.4$.

### 3 Details of the Data Applications

#### 3.1 Data Analyses of Autism Spectrum Disorder (ASD) in the SPARK Study

##### 3.1.1 Genotype Data Preprocessing and PGS Construction

We analyzed case-parent trio data from the Simons Foundation Powering Autism Research for Knowledge (SPARK) study[4]. The genotype phasing and imputation followed previous research[5]. Specifically, the imputation was performed on the Michigan imputation server using the Trans-Omics for Precision Medicine (TOPMed) Freeze 5b reference panel, which consisted of 125,568 haplotypes from multi-ancestry population. The SNPs with imputation quality $R^2 < 0.8$, missing call rates $> 1\%$, minor allele frequencies (MAF) $< 1\%$ were excluded.

The study population comprises 5 genetic ancestral groups determined using the HapMap3 reference panel: African (AFR), Americas (AMR), East Asian (EAS), European (EUR), and South Asian (SAS). The PGS scores were constructed with ambiguous SNPs removed, and their associated weights provided by recent external GWAS, as reported in the PGS catalog. To enable fair comparisons of estimated effect sizes across diverse ancestral populations, we further standardized the PGS values using 1000 Genomes (1000G) + HGDP unrelated individuals by constructing ASD PGS scores following the same procedures. In particular, these included 663 EUR, 684 AFR, 374 AMR, 718 EAS, and 671 SAS independent individuals from the 1000G + HGDP Project[6]. The PGS standardization details are described here[7]:

We first did mean adjustment through a linear regression of the raw PGS for each individual $i$ against the top five PCs derived from the PCA in 1000G + HGDP:

$$PGS_{i,1000G+HGDP} = \alpha_0 + \alpha_1 PC_{i1} + \cdots + \alpha_5 PC_{i5} + \epsilon_i$$

We then computed residuals $r_i$ of the raw PGS that account for mean differences in PGS distributions as

$$r_{i,1000G+HGDP} = PGS_{i,1000G+HGDP} - \hat{\alpha}_0 - \hat{\alpha}_1 PC_{i1,1000G+HGDP} - \cdots - \hat{\alpha}_5 PC_{i5,1000G+HGDP}$$

Finally, we take the regression coefficients and compute the ancestry-adjusted PGS for each individual $i$ in SPARK as

$$PGS_{j,standardized,ancestry} = \frac{PGS_{j,SPARK} - \hat{\alpha}_0 - \hat{\alpha}_1 PC_{j1,SPARK} - \cdots - \hat{\alpha}_5 PC_{j5,SPARK}}{SD(r_{1000G+HGDP,ancestry})}$$

Note that the denominator is the SD for residualized PGS with respect to each ancestry group in 1000G + HGDP. The reason we did the per-ancestry group standardization rather than using PC-projections in the denominator for each individual is to prevent the violation of Mendel's law within families, note that here, for each family, we assume that the ancestry group is defined the same as the offspring's ancestry group.

Variables we considered for the PGS x E interaction effects included maternal variables before pregnancy, including asthma, depression and other severe mental illness (defined as requiring medication or hospitalization), vitamin intake 3 months before pregnancy; variables during pregnancy including whether the mother experienced fever, eclampsia and preeclampsia, gestational diabetes, hyperemesis, pre-term or early labor; variables both before and during pregnancy including alcohol consumption and frequency, and smoking status; mother's age at birth, mother's educational attainment levels, and whether the child had low birth weight (defined as $< 2.5$kg). Mother's pregnancy period was defined as from 3 months before pregnancy to the end of breastfeeding.

For the PGS x context interaction, we used all 18 383 ASD case-parent trios. Genetic distance [8] is defined as the Euclidean distance between top 5 genetic PCs between each ASD-child and the center of 98 unrelated Finnish individuals from 1kGP+HGDP to represent the PGS training population (North European) from the iPSYCH consortium. We further calculated the genetic distance based on top 2 genetic PCs and top 10 PCs, the results for PGS x context interaction remain significant (Table S5).

We further used our model and the individual-level genotype data of ASD case-parent trios to estimate the ASD risk associated with several common polygenic predictors of cognitive-related traits and diseases, including education attainment[9], schizophrenia[10], strictly defined lifetime major depressive disorder[11], bipolar disorder[12], neuroticism[9], sleeplessness/insomnia[9], and attention-deficit/hyperactivity disorder (ADHD)[13]. As a negative control, we used body mass index (BMI)[9]. We constructed PGS using reported GWAS summary statistics of these traits, following the same procedures as described above. In particular, for ADHD, we used PRScs[14] and GWAS summary statistics following the steps reported previously[13] to construct PGS in our study. We standardized the PGS scores similarly using PC-projections as the ASD-PGS as described above for fair comparisons.

#### 3.2 Data Analyses of Non-Syndromic Orofacial Clefts (OFCs) in the GENEVA Study

##### 3.2.1 Genotype Data Preprocessing and PGS Construction

We investigated OFCs using case-parent trio data from the Gene Environment Association Studies initiative (GENEVA)[15]. GENEVA is a multi-ethnic study with data collected from Europe (Norway), the United States, and Asia (China, South Korea, Singapore, and the Philippines). Detailed genotype data imputation and quality control steps are described in previous work[16]. We additionally excluded SNPs with MAF $< 1\%$ and missing call rates $> 1\%$. After eliminating ambiguous SNPs, PGS values associated with cleft lip with or without cleft palate (CL/P) were computed using 24 SNPs and their respective weights sourced from the PGS catalog. These weights were based on summary statistics derived from multiple preceding GWAS studies. Note that due to the restricted data resources available in previous GWAS efforts, certain SNPs uncovered in specific studies either conducted meta-analyses or used data that partially intersected with samples in the GENEVA study. Nonetheless, these SNPs underwent subsequent validation using separate and independent data sources. We standardized the PGS values within each specific ancestry group using 503 EAS and 498 EUR independent individuals from the 1000G project.

For PGSxE interaction analysis, the maternal environmental exposures were collected through maternal interviews focused on the period from 3 months before pregnancy through the first trimester, which includes the first 8-9 weeks of gestation when palatal development is completed. The difference between maternal and paternal indirect PGS effects was also analyzed using our model. In the end, our analysis incorporated independent and complete 575 self-reported EUR and 891 Asian ancestry CL/P case-parent trios, and 203 EUR and 235 Asian CP case-parent trios.

#### 3.3 Association Studies of Genetically Predicted Multi-Omics Data on ASD and OFCs

We first built genetic scores for 12 539 whole blood gene expression levels and 140 serum metabolomic traits using summary statistics reported in OMICSPRED[17]. We then excluded biomolecular traits with variance explained $R^2 < 0.1$ by the genetic score in the internal validation or traits that contain fewer than 5 SNPs in the genetic score reported by OMICSPRED. So in the end we analyzed 27 metabolomics traits, and 4 907 genes for ASD and 4 991 genes for OFCs using PGS-TRI. All omics summary statistics were trained based on the INTERVAL EUR healthy blood cohort, and validated using multiple independent studies consisting of multi-ancestry populations. Specifically, the RNAseq summary statistics were trained based on the Illumina RNAseq platform using 4 136 individuals, and metabolomics summary statistics were based on the Nightingale platform trained using 37 359 individuals. Subsequently, we used PGS-TRI to conduct transcriptome-wide association studies and metabolome-wide association studies respectively to understand the potential molecular causal effects on ASD and OFCs risks using data from case-parent trios in SPARK and GENEVA studies. For each omics data type in each disease association study, we used the Benjamini-Hochberg false discovery rate (FDR) of 5% for multiple hypotheses testing adjustments.

We further investigated *CADM2* transcriptomic scores in brain tissues using precomputed genetic reference weights in GTEx v8[18]. We computed the scores using genes that achieved significant heritability and weights from "All Samples". Table S9 shows the results of the direct effect estimates for *CADM2* expression in brain tissues associated with ASD.
